# Triangulating evidence in health sciences with Annotated Semantic Queries

**DOI:** 10.1101/2022.04.12.22273803

**Authors:** Yi Liu, Tom R Gaunt

**Affiliations:** MRC Integrative Epidemiology Unit, Bristol Medical School, University of Bristol, Bristol, UK; NIHR Bristol Biomedical Research Centre, University of Bristol, Bristol, UK

## Abstract

Integrating information from data sources representing different study designs has the potential to strengthen evidence in population health research. However, this concept of evidence “triangulation” presents a number of challenges for systematically identifying and integrating relevant information. We present ASQ (Annotated Semantic Queries), a natural language query interface to the integrated biomedical entities and epidemiological evidence in EpiGraphDB, which enables users to extract “claims” from a piece of unstructured text, and then investigate the evidence that could either support, contradict the claims, or offer additional information to the query. This approach has the potential to support the rapid review of pre-prints, grant applications, conference abstracts and articles submitted for peer review. ASQ implements strategies to harmonize biomedical entities in different taxonomies and evidence from different sources, to facilitate evidence triangulation and interpretation. ASQ is openly available at https://asq.epigraphdb.org.

## 1 Introduction

Researchers in health sciences are encouraged to seek multiple strands of complementary evidence to minimise the risk of bias creating false positives. This has been referred to as the *triangulation*^1^ of evidence, which may combine results from different study designs with different sources of bias, including from established findings in the literature. Platforms which offer a portal to integrated heterogeneous data such as Open Targets^2^ and EpiGraphDB^3^ are highly valuable sources which have the potential to support evidence triangulation by integrating evidence with relevant information from a range of dedicated data providers, including biomedical ontologies^4 5^, genetic associations^6^ and literature-derived evidence^7^. One of the main objectives for the web interface of such integrated data platforms is to present users with focused information from various integrated sources in order to facilitate the fast navigation and discovery of evidence. However, there is a need to improve accessibility of such complex data resources for less experienced users and to improve the interpretability of data, transforming source data into comprehensible evidence and knowledge regarding a research question. There are several challenges in order for these issues to be addressed, such as: how can a research question be represented so that evidence can be retrieved for triangulation, how should we integrate biomedical entities from different taxonomies and their relationships, and what strategies should we use to form larger *evidence* groups from heterogeneous source data and methods to prioritise the retrieved evidence?

We approach the above-mentioned challenges by developing a scientific claim query platform Annotated Semantic Queries (ASQ; https://asq.epigraphdb.org) on top of the integrated biomedical knowledge and evidence of the EpiGraphDB^3^ data and ecosystem, where users are able to investigate the various groups of evidence that support / contradict a claim, and assist their further investigation into the source data that relates to that claim. Scientific claims from a query text (e.g. the abstract of a pre-print) are parsed into claim triples (in the form of `Subject PREDICATE Object`) in ASQ, where the subject and object terms are annotated with ontology terms and then mapped to evidence entities in an *entity harmonization* process, by a combination of high-dimensional text embeddings and sequence classification methods. Evidence from difference sources are then retrieved and harmonized into two groups – a *triple and literature* evidence group comprised of literature and literature-derived evidence, and an *association* evidence group comprised of statistical association results from systematic analyses. Evidence items are then assigned with scores that reflect their strength as well as their relevance to the claim, so that ASQ is able to present evidence which would be of potential high value to the user in order for them to assess and triangulate the evidence regarding a scientific claim. Here we discuss the implementation of the ASQ platform and components and demonstrate its use for systematic analysis of claims derived from MedRxiv^8^ submissions from 2020 to 2021.

## 2 Results

### 2.1 The EpiGraphDB-ASQ platform

The ASQ platform is developed as a natural language interface component to the epidemiological evidence integrated in EpiGraphDB database and ecosystem (Figure 1), with the aim of allowing users to access EpiGraphDB knowledge and triangulate the evidence using a simple scientific claim of interest as a starting point. For example, instead of relying on bespoke topic-specific web queries that are restricted to several entities or meta-entities or via structural queries to the database, ASQ presents the integrated evidence from EpiGraphDB as introspectable evidence items that “fact-check” a claim “glucose can be used to treat diabetes” (Figure 2) or a short piece of text containing multiple such claims.

**Figure 1.**
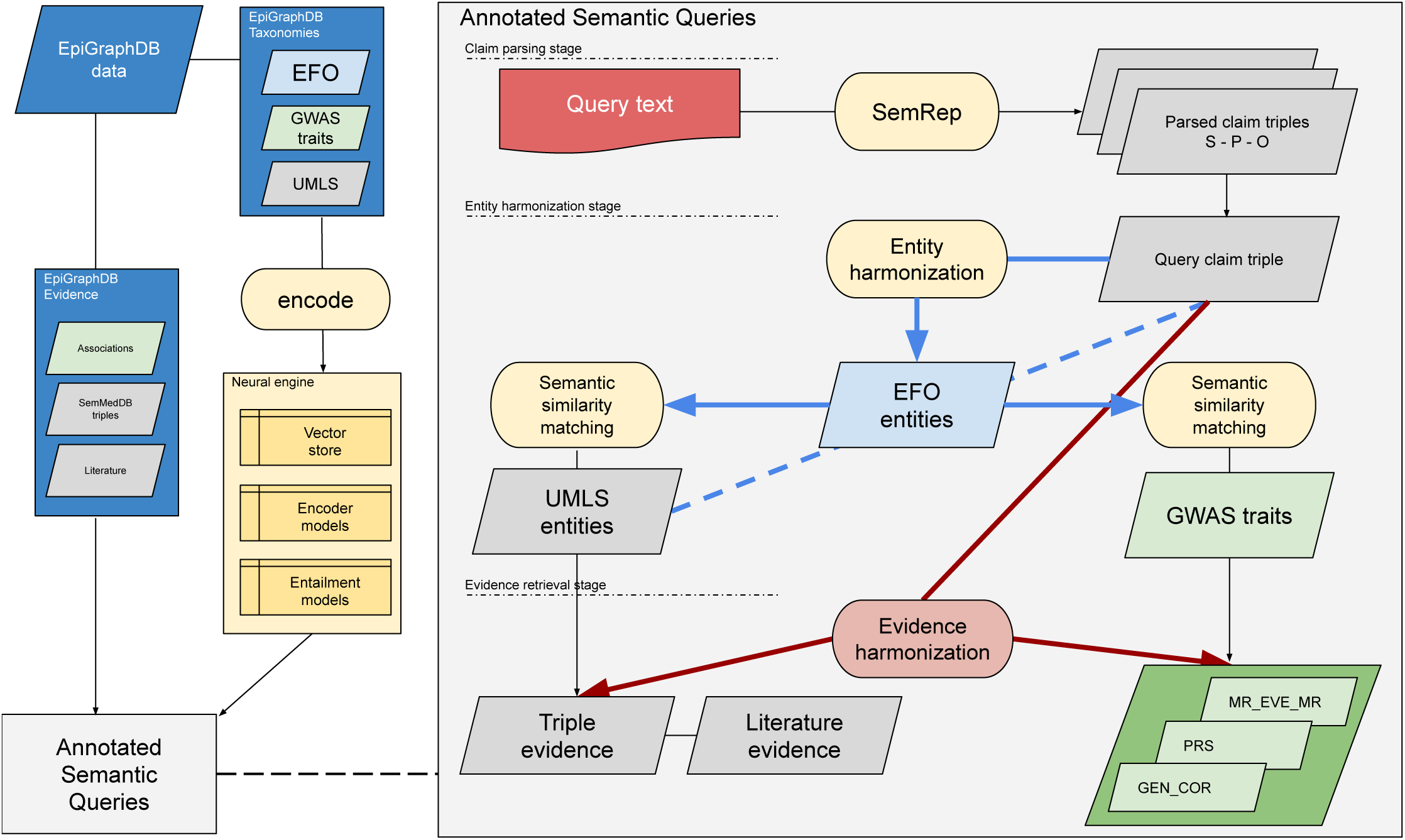
Architecture of the EpiGraphDB-ASQ platform. Overall architecture design of the EpiGraphDB-ASQ platform and its associated components in the EpiGraphDB ecosystem. **Left**: EpiGraphDB’s biomedical entities (in the form of graph nodes) from different taxonomies are encoded into vector representations which allows for fast information retrieval against the query of interest. Epidemiological evidence (in the form of graph edges) are incorporated into ASQ as harmonized evidence groups. **Right**: Internal processing workflow of the EpiGraphDB-ASQ platform by the three stages: the claim parsing stage, the entity harmonization stage, and the evidence retrieval stage.

**Figure 2.**
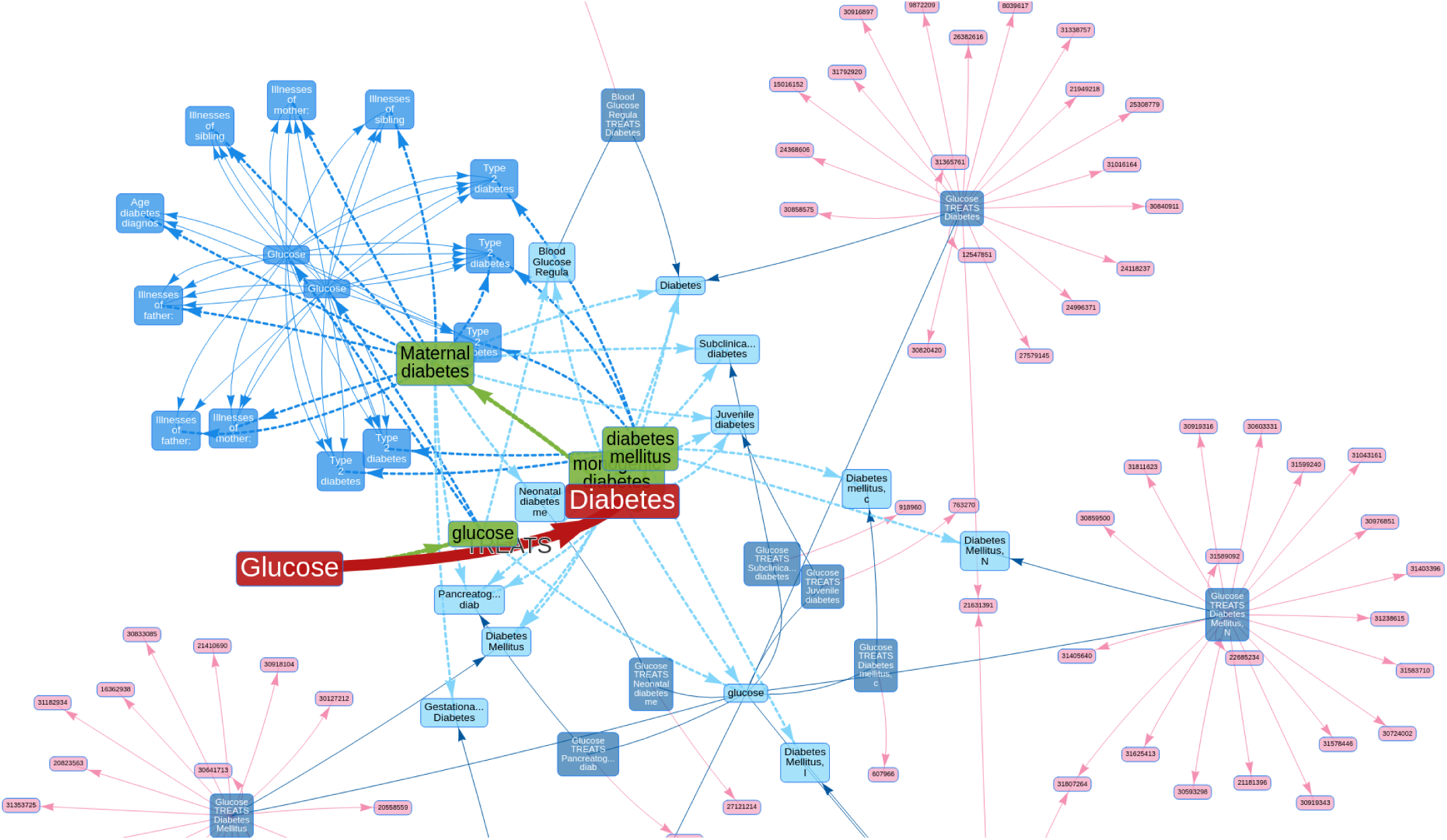
Evidence triangulation regarding scientific claims. A summary network diagram on the retrieved entities and evidence from the ASQ platform regarding a claim “Glucose TREATS Diabetes”. The subject and object terms of the query claim are represented as nodes in red, and the predicate as a directed edge. The ontology term (green nodes) “glucose” is identified as the mapped term for the claim subject, and ontology terms “diabetes mellitus”, “monogenic diabetes”, “Maternal diabetes” are identified as the mapped terms for the claim object in the default setting (which can be adjusted at an interactive session or updated after initial results). Triple and literature evidence are represented as semantic triples (deep blue nodes) formed by UMLS Metathesaurus terms (light blue nodes), which are linked to source literature findings (pink nodes). For association evidence, statistical association results on GWAS traits (blue nodes) are represented as edges between them. Edges in dashed lines represent mappings between taxonomies, and edges in solid lines represent evidence items.

Various components of epidemiological evidence from EpiGraphDB are incorporated into the ASQ platform as two **evidence groups** (Table 1):

1. A *triple and literature* evidence group which consists of both the semantic triples derived from literature sources that are integrated from SemMedDB^7^ and the source literature articles from PubMed from which the triples are derived (Supplementary tables 1 and 2). Typically an evidence item in this group is comprised of a semantic triple in the form of `Subject-PREDICATE-Object` (e.g. “Obesity CAUSES Asthma”) and the multiple source literature items containing specific context details in the literature title and abstract text. SemMedDB imports parsed entities and triples from SemRep^9^ which normalizes terms mentioned in a text document into UMLS Metathesaurus entities^10^ and predicate relationships into Semantic Network relationships^11^.
2. An *association* evidence group which consists of various sources of curated systematic statistical association analysis studies using systematic Mendelian Randomization analyses (the `[MR_EVE_MR]` ^12^ relationships in EpiGraphDB data; see Supplementary Table 4 for notation conventions), genetic correlations (the `[GEN_COR]` ^13^ relationships), and polygenic risk score associations (the `[PRS]` ^14^ relationships), where the analyses are conducted between two human traits for which genome-wide association study (GWAS) data are curated by OpenGWAS^6^ (the `(Gwas)` nodes in EpiGraphDB). ASQ incorporates the common properties of *effect size, standard error, P-Value*, as well as *source*/*target* GWAS traits from the source analysis data as the common quantitative/qualitative information of the evidence items, and additional detailed source-specific properties are also retrieved for users’ own investigation.

**Table 1.** Integration of epidemiological evidence in EpiGraphDB. Distribution of the EpiGraphDB *knowledge* triples which are the *source* evidence in this study, harmonized into the two evidence categories. Column “Triples” and column “Literature” report respectively the number of literature triples and number of associated source literature articles in a triple and literature evidence group, and column “Associations” report the number of statistical associations in an association evidence group. For example, there are 37,423 literature triples in the form of `Term 1 AFFECTS Term 2` where `Term 1` and `Term 2` are from the term types of `aapp` (“Amino Acid, Peptide, or Protein”), `dsyn` (“Disease or Syndrome”), `gngm` (“Gene or Genome”) (a UMLS term can have multiple associated types), and there are 57,928 source literature articles from which the 37,423 literature triples are derived. Similarly, there are 8,966,440 statistical associations from the MR-EvE study^12^ between GWAS-es in the UKBiobank categories (`ukb-a`, `ukb-b`, etc.) of OpenGWAS^6^. A UMLS term can have multiple associated semantic types and the label descriptions on UMLS semantic types are available in Supplementary Table 1.

| Triple and literature evidence |  |  |  |  |
| --- | --- | --- | --- | --- |
| Direction | UMLS Predicate | UMLS term type | Triples | Literature |
| Directional | AFFECTS | *aapp*, *dsyn*, *gngm* | 37,243 | 57,928 |
|  | AFFECTS | *dsyn* | 29,167 | 58,753 |
|  | CAUSES | *dsyn* | 85,231 | 222,462 |
|  | CAUSES | *aapp*, *dsyn*, *gngm* | 49,178 | 100,681 |
|  | TREATS | *phsu*, *dsyn*, *orch* | 82,263 | 274,589 |
|  | TREATS | *phsu*, *dsyn* | 47,416 | 238,636 |
|  | PRODUCES | *aapp*, *gngm* | 69,691 | 106,862 |
|  | PRODUCES | *phsu*, *aapp*, *gngm* | 12,706 | 26,122 |
| Non-directional | ASSOCIATED_WITH | *aapp*, *dsyn*, *gngm* | 188,961 | 423,727 |
|  | ASSOCIATED_WITH | *phsu*, *aapp*, *dsyn*, *gngm* | 29,425 | 86,176 |
|  | INTERACTS_WITH | *aapp*, *gngm* | 393,759 | 673,470 |
|  | COEXISTS_WITH | *aapp*, *gngm* | 224,098 | 332,834 |
|  | COEXISTS_WITH | *dsyn* | 150,166 | 385,349 |
|  | INTERACTS_WITH | *aapp*, *enzy*, *gngm* | 72,194 | 140,836 |
| Association evidence |  |  |  |  |
| Direction | Association type | GWAS categories | Associations |  |
| Directional | MR_EVE_MR | *ukb*, *ukb* | 8,966,440 |  |
|  | MR_EVE_MR | *prot*, *ukb* | 5,028,904 |  |
|  | MR_EVE_MR | *ubm*, *ukb* | 3,833,948 |  |
|  | MR_EVE_MR | *prot*, *prot* | 3,109,406 |  |
|  | MR_EVE_MR | *prot*, *ubm* | 1,974,611 |  |
| Non-directional | GEN_COR | *ukb-b*, *ukb-b* | 453,752 |  |
|  | GEN_COR | *ukb-a*, *ukb-b* | 286,536 |  |
|  | GEN_COR | *ukb-a*, *ukb-a* | 180,536 |  |
|  | GEN_COR | *ukb-b*, *ukb-d* | 133,554 |  |
|  | GEN_COR | *ukb-a*, *ukb-d* | 84,266 |  |
|  | GEN_COR | *ukb-d*, *ukb-d* | 38,908 |  |
|  | PRS | *ieu-a*, *ukb-a* | 70,926 |  |
|  | PRS | *ukb-b*, *ieu-a* | 45,394 |  |
|  | PRS | *ukb-a*, *ukb-a* | 2,198 |  |
|  | PRS | *ukb-b*, *ukb-a* | 704 |  |

On the web interface, the main entry point for a user to interact with the platform is to input short paragraphs of scientific text (e.g. the abstract of a journal article or pre-print). From this input we use SemRep^9^ as the query parser to derive **query claim triples** from the text in the form of `Subject-PREDICATE-Object` (e.g. “Obesity CAUSES Asthma”). The user is then asked to select a specific triple of interest as the target of the downstream stages of entity harmonization and evidence retrieval. Alternatively, users can either directly input a query claim in the query triple view (https://asq.epigraphdb.org/triple), or start from the MedRxiv systematic analysis summary results (https://asq.epigraphdb.org/medrxiv-analysis).

In the following *entity harmonization* stage, ASQ harmonizes the biomedical entities from the claim triple with the Experimental Factor Ontology (EFO^5^) entities, with the EFO ontology serving as the anchor to connect the query entities and any evidence entities (Section 4.1). By default ASQ attempts to retrieve entities that are semantically highly related (but not exclusively identical) to the query entities to allow for exploratory discovery about further evidence of potential interest. This can be adjusted to more restrictive (specific) or more liberal (sensitive) mapping.

In the *evidence retrieval* stage, evidence items from the two evidence groups are retrieved based on the biomedical entities harmonized in the previous stage, as well as on the **predicate direction group** (“directional” and “non-directional”) of the claim triple. Evidence items are then categorised into several **evidence types** (Section 4.2): (a) supports the query claim (“supporting”), (b) contradicts with the query claim with retrieved items indicating evidence in the opposite direction to the claim (“reversal”), (c) fail to meet the required evidence threshold to be supporting or contradictory (“insufficient”) or (d) could be of additional information (“additional”) to the claim. Retrieved individual evidence item and groups of items are then measured with a score to reflect both the proximity of the involved entity to the query claim as well as the strength of the evidence (Section 4.3). For triple and literature evidence the strength of the evidence item is calculated based on the number of literature sources, whereas for association evidence the strength is calculated based on the standardized effect size. The evidence strength score is then adjusted by a mapping score measuring the semantic similarities between the subject/object terms of the evidence item and those of the query claim.

ASQ provides comprehensive information (Figure 3) regarding the status of entity harmonization with summary diagrams describing the mapped ontology entities and harmonization metrics linking the evidence entities to the original query claim. For triple and literature evidence, users can further introspect the context detail from which the semantic triples are derived, and for association evidence, ASQ displays the statistical results on forest plots as quantitative comparisons. In addition to the default interactive session on the web interface, ASQ offers programmatic access via the API (See “Code availability” section) which allows for batch processing and analysis (e.g. Section 2.2).

**Figure 3.**
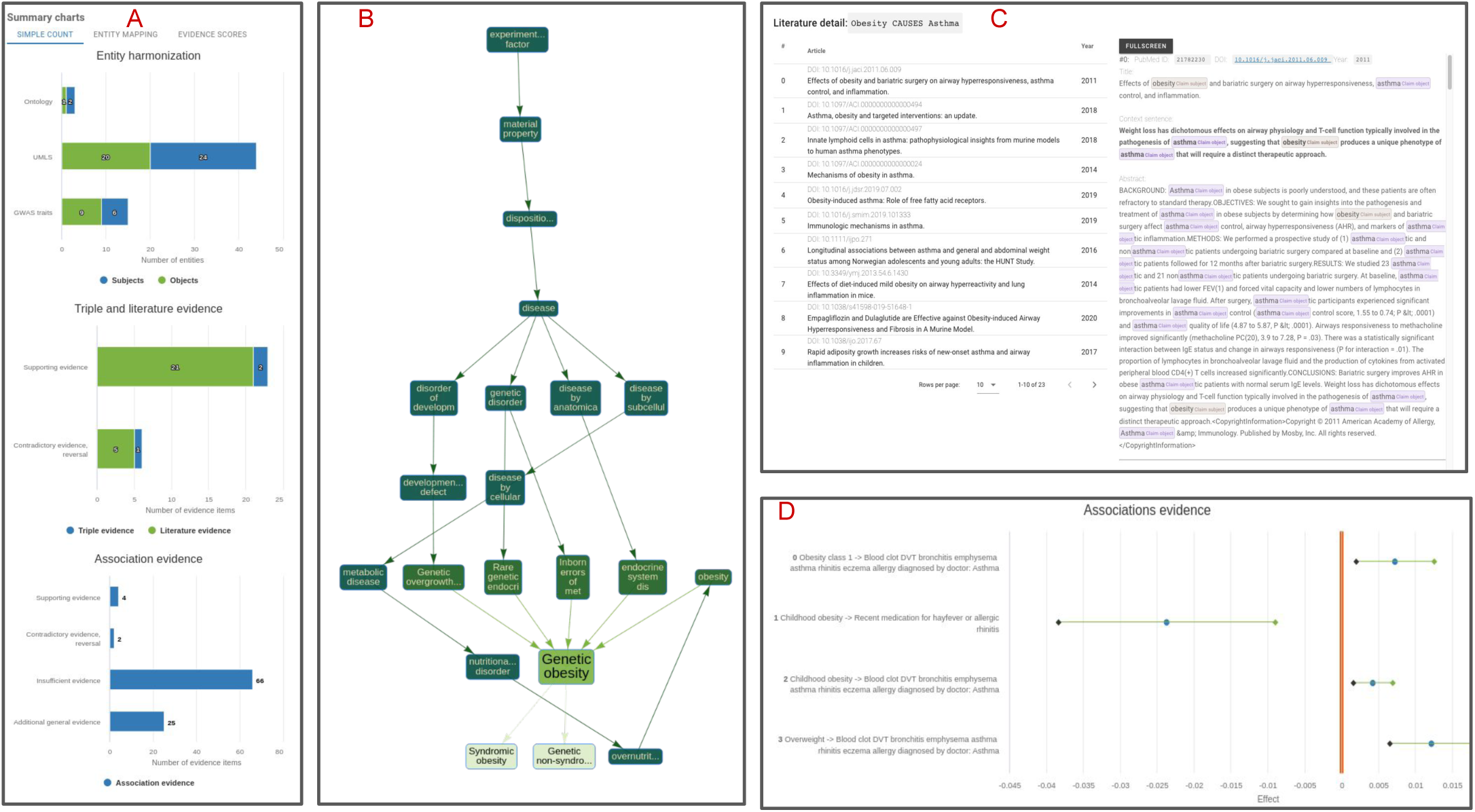
Overview of web interface. Overview of the web interface functionalities of EpiGraphDB-ASQ. **A**: Summary of harmonized entities and retrieved evidence regarding the query claim. **B**: Sub-graph representation of a retrieved ontology entity in the EFO graph. **C**: Summarised literature information and context details for a retrieved semantic triple “Obesity CAUSES Asthma”. **D**: Forest plot on the statistical association evidence regarding the query claim.

### 2.2 Systematic analysis of MedRxiv submissions

#### 2.2.1 Study design

We demonstrate the use of ASQ by systematically analysing the preprint submissions on MedRxiv in the sample period from 2020-01-01 to 2021-12-31 (Figure 4). We will further discuss the technical details and the relevant terminology covered here in Section 4. Using the MedRxiv/BioRxiv API, we identified 28,846 unique submissions in the period (in the case of multiple versions in a submission we kept only the initial version) and retrieved their abstracts as candidate text documents containing multiple scientific claims to be parsed in SemRep. Out of all the candidate documents, 13,999 documents were successfully parsed by SemRep to contain coherent semantic triples at sentence level, and 6,870 documents were identified to contain suitable predicates for analysis in ASQ. In total we extracted 13,295 document-triples (14,436 claim triples) as the sample dataset.

**Figure 4.**
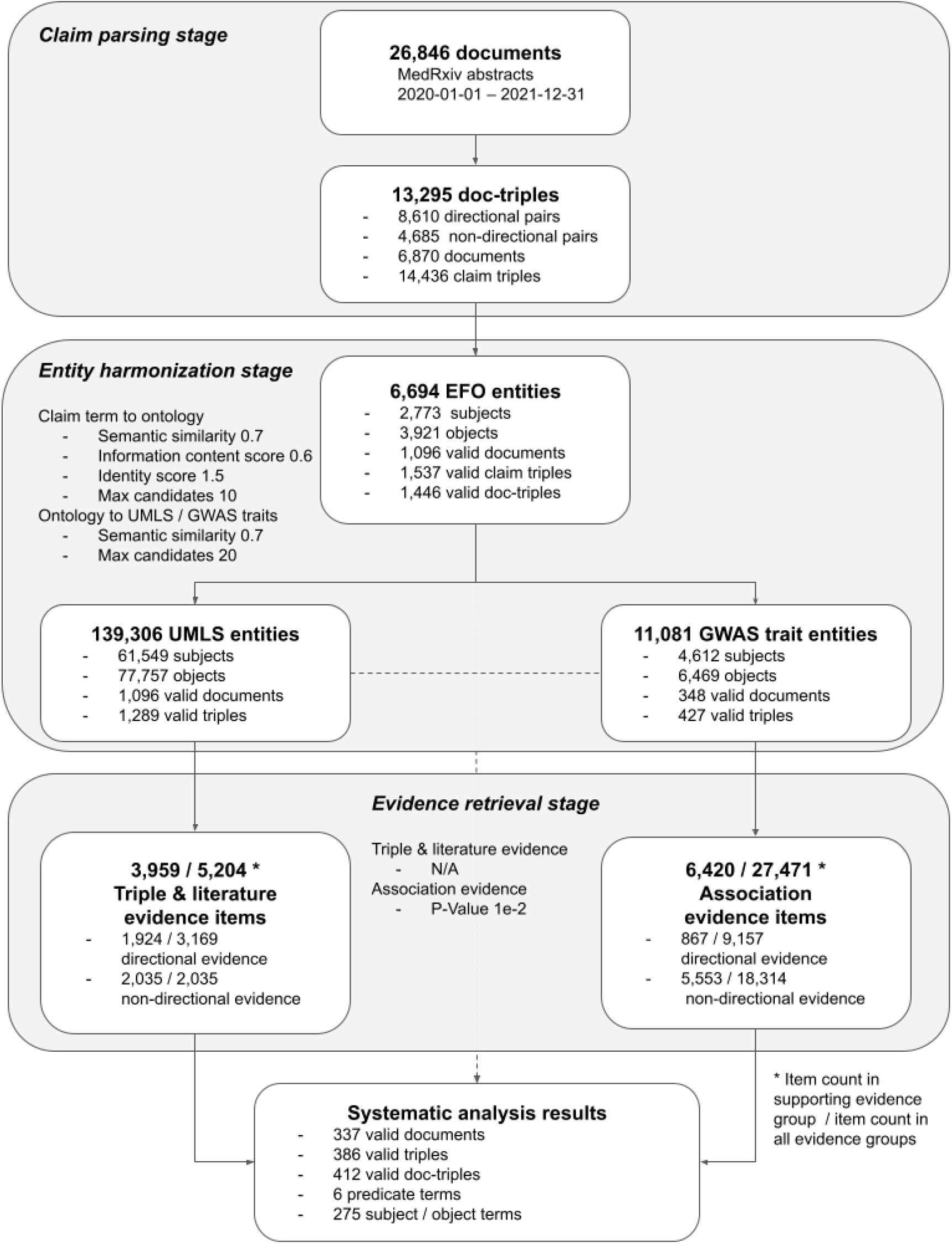
Study design of the systematic analysis and result metrics. Overview diagram on the systematic analysis results and the primary metrics in the various stages discussed in Section 2.2. This figure complements Figure 1 regarding an individual case with the aspect of systematic scale. Further discussions on the parameter configuration as shown in each of the stages are available in Supplementary Table 5.

Each claim triple was processed through ASQ programmatically to map with EFO and evidence entities in the entity harmonization stage using a set of parameters which are equivalent to the default settings used in the web interface (see Supplementary Table 5 for configuration of parameters), with 1,446 document-triples identified to be valid and associated with entities in EpiGraphDB. Amongst these document-triples, we found that “Disease or Syndrome” is the most numerous semantic group (888 query terms, 7,831 EFO entities), followed by “Mental or Behavioral Dysfunction” (125 query terms, 1,708 EFO entities), and “Neoplastic Process” (138 query terms, 1,516 EFO entities) (Supplementary Table 7). In order to avoid the document-triple dataset for analysis being too large we used the intersection subset where a document-triple must contain at least one type of evidence in both the *triple and literature evidence* group and the *association evidence* group (Supplementary Table 6 reports the summary statistics). The final dataset consisted of 412 claim triples (337 submission abstracts, and 386 document-triples) where each claim is associated with retrieved evidence from multiple evidence types across the evidence groups, and can be accessed via https://asq.epigraphdb.org/medrxiv-analysis with the option to adjust settings for individual query cases.

#### 2.2.2 Systematic results from entity harmonization and evidence retrieval

In the entity harmonization stage of the systematic analysis, the retrieval of EFO entities is determined by an initial stage where EFO candidates are retrieved by the semantic similarities between the EFO candidates and the query subject/object terms by their encoded text vectors, and a subset is subsequently selected based on proximity of the query term and the candidates in the EFO graph as indicated by the identity scores, which then gets mapped to evidence entities via semantic similarities (See Section 4.1). Figure 5 shows the distribution of score metrics for the entity harmonization process where query terms are mapped to EFO entities and Supplementary Figure 1 shows the distribution of scores for mappings of evidence entities to the original query terms. For selected entities which have identity scores below the threshold in the automated process, they would also be semantically closer to the query terms than the rest of the retrieved candidates (with mean semantic similarity scores above 0.9), and therefore from a systematic scale ASQ is able to select a set of corresponding EFO entities that have high association to the query terms of interest as the basis for further retrieving evidence entities related to these query terms. Similar automated approach applies in the interactive session, and users are able to optionally override the automated processing of entity harmonization with manual selection of EFO entities of interest or re-adjust the entity selection afterwards.

**Figure 5.**
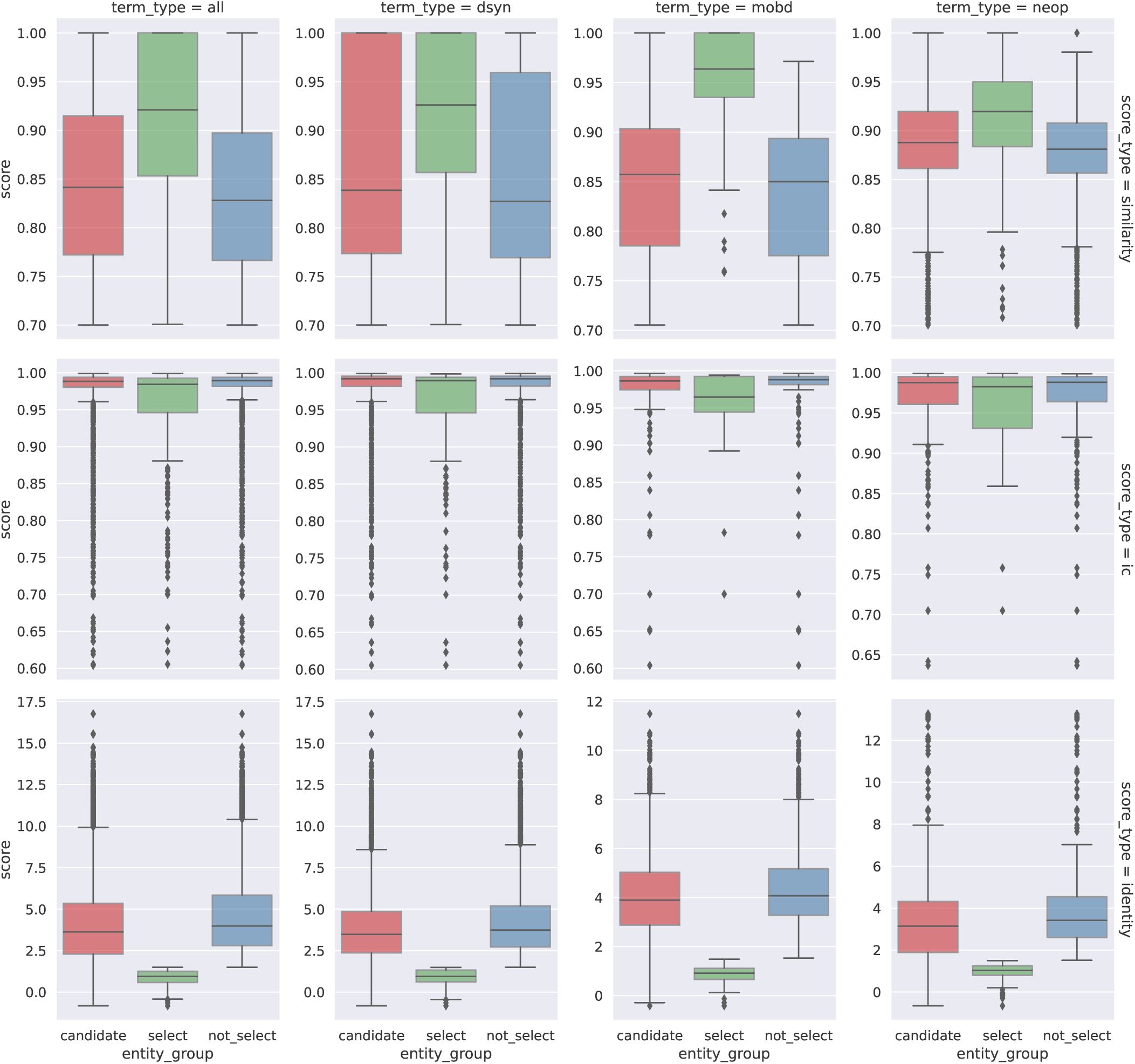
Entity harmonization stage: distribution of score metrics of retrieved EFO entities. Distribution of semantic similarity scores, information content scores, and identity scores for retrieved EFO entities in the process of mapping with query UMLS terms, categorised by the semantic type of the UMLS term (“term_type”) and score metrics (“score_type”) in the retrieval process. **Category type**: “candidate” for entities retrieved as a candidate, “select” for candidates that are selected in the automated process (Section 4.1), and “not_select” for candidates that are not selected. **Left** (“all”): Distribution across all semantic types. **Middle 1** (“dsyn”): In the “Disease or Syndrome” group. **Middle 2** (“mobd”): In the “Mental or Behavioral Dysfunction” group. **Right** (“neop”): In the “Neoplastic Process” group. This figure reports distributions in the top 3 semantic type groups by entity count (Supplementary Table 7 reports entity counts of all semantic types). **Top** (“similarity”): By semantic similarity score to measure similarity of term embedding vectors. **Center** (“ic”): By information content score to measure the concreteness of the term in EFO. **Bottom** (“identity”): By identity score to measure the inferred relative distance of the UMLS term. The roles of the score metrics take in the harmonization retrieval process are discussed in detail in Section 4.1. Supplementary figure 1 reports the distribution of score metrics for retrieved UMLS and trait entities.

Figure 6 shows the distribution of the evidence scores and their constituent scores in the evidence retrieval process, and further details are available in the supplementary materials on group specific distributions (Supplementary Figures 4 and 5), as well as on summary statistics (Supplementary Table 8). The mean mapping score for retrieved evidence is around 0.5 to 0.7, with a typical scenario where the retrieved entities have about 0.85 to 0.92 in semantic similarity to its upstream entities. The mean strength score for *triple and literature evidence* is around 1, i.e. on average a triple evidence item was identified in only 1 source literature item, though there is a long upper tail with many cases of 10 to 20 source literature items associated with a triple (i.e. strength scores around 2 to 2.3). Association evidence items are required to have sufficient strength of effect size in order to qualify into the “supporting” type or the “reversal” type (otherwise they would be of “insufficient” evidence), and therefore the strength scores for “supporting” and “reversal” types are markedly higher than items in the “insufficient” type. In general, the evidence scores for “supporting” and “reversal” *association evidence* are found to be distributed around the baseline score of 1. In addition, as constituent scores the mapping scores and strength scores contribute to the evidence scores in roughly linear relationships (Supplementary Figures 2 and 3).

**Figure 6.**
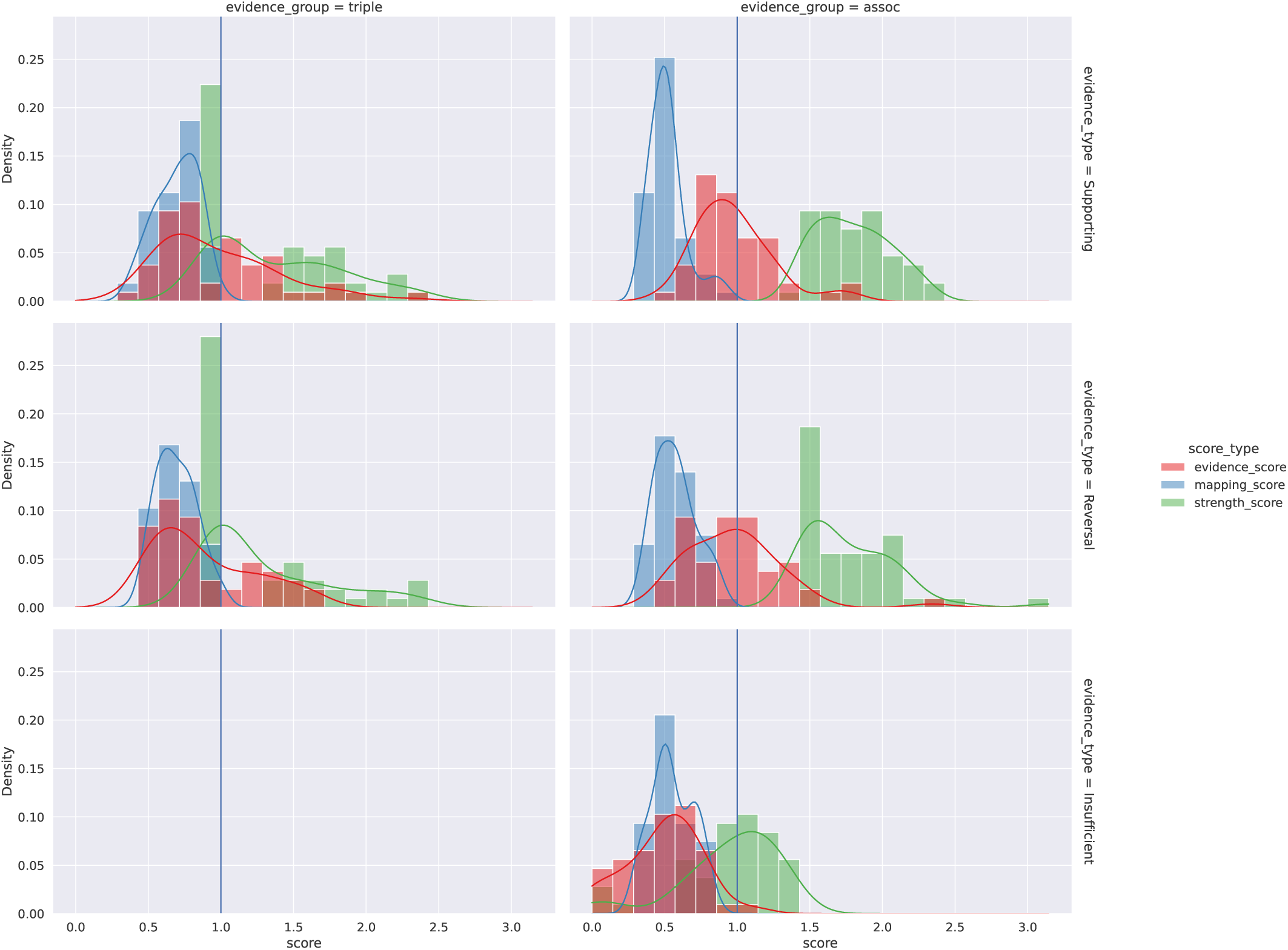
Evidence retrieval stage: distribution of evidence scores and constituent scores. Distribution of evidence scores and its constituent scores (entity mapping scores and evidence strength scores), for the “supporting”, “reversal”, and “insufficient” evidence types (by rows) in the triple and literature evidence group and the association evidence group (by columns). This figure reports aggregated distributions across directional and non-directional predicate groups, and Supplementary Tables 4 and 5 report detailed distributions by evidence groups, evidence types, and predicate groups. Note an “insufficient” evidence type is only applicable to the association (“assoc”) evidence group and not the triple and literature (“triple”) evidence group.

#### 2.2.3 Top cases of research areas

Whilst results from the systematic analysis reflect the availability of evidence in ASQ and EpiGraphDB in various areas, they also show the popular research topics and themes reflected from MedRxiv submissions in 2020-2021. Figure 7 shows several clusters of research areas with central terms as measured in Table 2, and example claim triples can be found in Supplementary Table 9. In addition to research associated with the COVID-19 pandemic (“Coronavirus infections”), the two areas with highest research submissions and retrieved evidence are regarding obesity and associated diseases (“Obesity”, “Diabetes”, “Diabetes Mellitus, Non-Insulin-Dependent”, “Chronic Kidney Diseases”, etc.) and mental health (“Depressive disorder”, “Parkinson Disease”, “Alzheimer’s Disease”, “Schizophrenia”, etc.). Notably when SemRep fails to recognize a more specific term it will fall back to more general terms, and therefore the term “Disease” is prominent in the list of top terms. Examples in Supplementary Table 9 suggest claims involving predicates “CAUSES”, “AFFECTS” and “COEXISTS_WITH” are the most popular claims that can be derived from submitted abstracts, which is similar to the summarised results in all cases in Supplementary Table 8 where these predicates are the ones with most retrieved evidence items.

**Figure 7.**
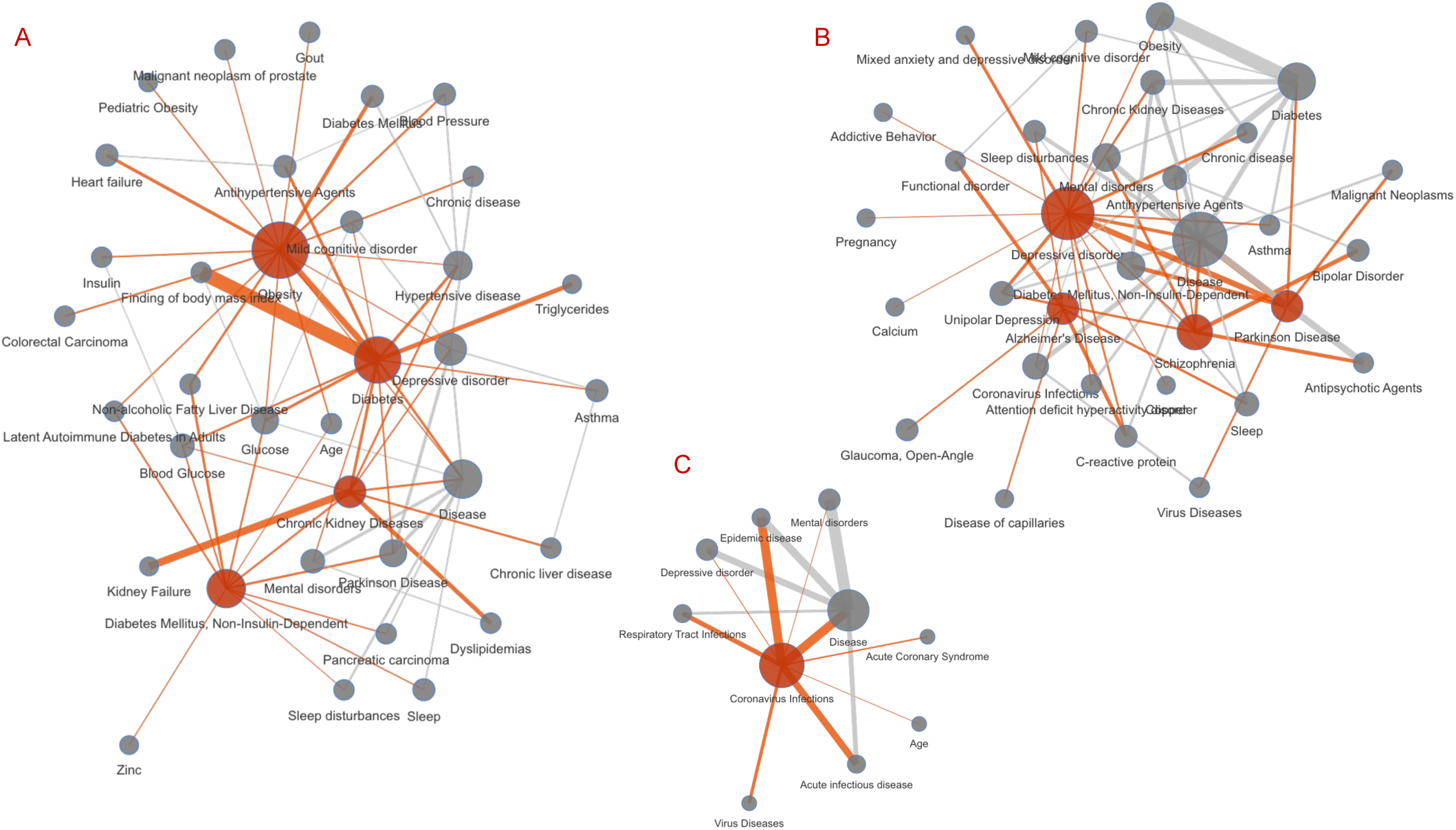
Systematic analysis results: evidence clusters of topic terms. Clusters representing research interests as parsed from the MedRxiv abstract sample from 2020-01-01 to 2021-12-31 as well as their corresponding evidence retrieved from EpiGraphDB-ASQ as network diagrams. Nodes coloured in red correspond to the primary claim terms (Table 2) and edges coloured in red correspond to relationships involving a primary claim term. **A**: Obesity cluster with primary terms “Obesity”, “Diabetes”, “Diabetes Mellitus, Non-Insulin-Dependent”, “Chronic Kidney Diseases”; **B**: Mental illness cluster with primary terms “Depressive disorder”, “Alzheimer’s Disease”, “Schizophrenia”, “Parkinson Disease”; **C**: COVID-19 cluster with primary terms “Coronavirus infections”; The diagrams are generated by retrieving first-degree neighbour nodes for each of the top term nodes, where node size corresponds to term count, and edge width correspond to aggregated supporting evidence scores between nodes. Interactive diagram is available on https://asq.epigraphdb.org/medrxiv-analysis.

**Table 2.** Systematic analysis results: top claim terms by retrieved evidence. Top claim terms sorted by the number of cases where the query claim triple is associated with both triple and literature evidence as well as association evidence (“T&L. + Assoc.”). For example, there are 41 claim triples involving the term “Disease” as either a subject term or an object term where these claim triples are identified with *both* supporting evidence in triple and literature evidence (“T&L.”) and association evidence (“Assoc.”) groups, 74 cases identified with supporting triple evidence, 44 cases identified with supporting association evidence, 77 cases identified with any evidence types (“Any”; See Section 4.2 and Table 3 for all evidence types), and 715 cases from the triples in the initial claim parsing stage dataset (“Init.”; Figure 4) after parsing from source abstracts regardless of results in the downstream stages.

| Claim term | Supporting |  |  | Any | Init. |
| --- | --- | --- | --- | --- | --- |
|  | T&L. + Assoc. | T&L. | Assoc. |  |  |
| Disease | 41 | 74 | 44 | 77 | 715 |
| Obesity | 20 | 25 | 25 | 30 | 125 |
| Diabetes | 17 | 19 | 18 | 20 | 87 |
| Depressive disorder | 14 | 20 | 16 | 26 | 100 |
| Parkinson Disease | 13 | 13 | 13 | 13 | 111 |
| Diabetes Mellitus, Non-Insulin-Dependent | 10 | 12 | 12 | 15 | 84 |
| Alzheimer’s Disease | 8 | 10 | 8 | 10 | 111 |
| Schizophrenia | 8 | 11 | 8 | 11 | 32 |
| C-reactive protein | 7 | 7 | 9 | 10 | 24 |
| Malignant Neoplasms | 7 | 8 | 15 | 19 | 100 |
| Chronic Kidney Diseases | 6 | 9 | 6 | 9 | 35 |
| Chronic disease | 5 | 6 | 5 | 6 | 44 |
| Fatigue | 5 | 5 | 6 | 6 | 25 |
| Sleep | 5 | 5 | 6 | 6 | 21 |
| Atrial Fibrillation | 5 | 6 | 6 | 9 | 57 |
| Pain | 4 | 4 | 6 | 6 | 30 |
| Glucose | 4 | 5 | 4 | 6 | 20 |
| Blood Glucose | 4 | 5 | 4 | 5 | 15 |
| Hypertensive disease | 4 | 12 | 4 | 13 | 90 |
| Mental disorders | 4 | 8 | 4 | 10 | 42 |
| Cardioembolic stroke | 3 | 3 | 3 | 3 | 14 |
| Testosterone | 3 | 5 | 3 | 6 | 21 |
| Diabetes Mellitus | 3 | 4 | 5 | 6 | 25 |
| Triglycerides | 3 | 4 | 4 | 5 | 16 |
| Heart Diseases | 3 | 3 | 3 | 3 | 10 |
| Unipolar Depression | 3 | 4 | 3 | 4 | 32 |
| Myocardial Infarction | 3 | 4 | 5 | 6 | 15 |
| Malignant neoplasm of prostate | 3 | 4 | 3 | 4 | 18 |
| Enthesitis-Related Arthritis | 3 | 4 | 3 | 4 | 25 |
| Behavior | 3 | 3 | 3 | 4 | 13 |

#### 2.2.4 Individual case

Here we showcase an example in demonstrating the use of ASQ for researchers in triangulating evidence regarding epidemiological research questions. From the systematic dataset, ASQ extracted a claim triple “Obesity CAUSES Heart failure” from a preprint abstract regarding a Mendelian randomization analysis investigating causal relationships between body mass index and heart failure risk^15^, derived from the context “About 40% of the excess risk of HF due to adiposity is driven by SBP, AF, DM and CHD”, where “HF” is recognised as heart failure and “adiposity” as obesity. These results can be found on ASQ (https://asq.epigraphdb.org/triple?subject=Obesity&object=Heart%20failure&predicate=CAUSES&analysis). The query subject “Obesity” and object “Heart failure” were mapped to their corresponding ontology counterparts then to evidence entities, from which ASQ then identified suitable evidence items. At aggregate level for *triple and literature evidence* there are more supporting evidence items (11) with higher aggregated scores (12.80) compared to reversal evidence items (6) with lower aggregated scores (5.46), similarly there are 5 supporting *association evidence* items with an aggregated score of 5.11 with no reversal evidence identified. Users are able to further investigate the literature that either associate with the claim triple (e.g.^16^ and^17^) or the reversal claim that heart failure might cause obesity (e.g.^18^), viewing the surrounding context from the abstract directly in the ASQ interface, or clicking a link to access the original paper. For *association evidence* ASQ identified several individual findings from the pairwise Mendelian randomization studies with sufficient statistical significance as supporting evidence. ASQ also identified a range of findings that are insufficient in statistical significance to qualify as supporting evidence, which are useful both in showing the scope of evidence identification but also in determining the cause of a lack of reversal evidence. In this case, the lack of reversal evidence was due to absence of results from the MR-EvE data source (as there were no retrieved insufficient counterparts to reversal evidence items). In addition, ASQ identified several findings from the PRS Atlas data source, and since the identified trait term “Target heart rate achieved” was not directly equivalent to the query object “Heart failure” ASQ would assign low evidence scores to these findings in the context of the original claim. In general ASQ is able to assist researchers in investigating research questions in epidemiology both at aggregate level to have an overview of the evidence categories regarding the question, as well as at individual level for researchers to further investigate the evidence items in literature or statistical findings with their expert knowledge.

## 3 Discussions

We developed the Annotated Semantic Queries (ASQ) platform as an approach to improve the accessibility of the EpiGraphDB data and ecosystem for users through the implementation of a natural language interface (whilst also enhancing programmatic access). There is an intrinsic problem with integrated data platforms containing rich and complex data: experienced users wish to be presented with flexible access to the data in order to navigate to the elements they want, yet new users can find this complexity overwhelming (even if well documented). From this perspective ASQ provides an accessible natural language query interface for such users to find the evidence relating to a specific claim/question e.g. “Can obesity cause asthma?”, which can either be parsed from a short piece of text containing scientific claims, or directly input as a claim triple of `Subject PREDICATE Object`. In addition to providing a more accessible interface to EpiGraphDB this approach provides a novel way to systematically evaluate a piece of text (such as a pre-print abstract) to identify whether claims within that text are supported by other data. Heterogeneous knowledge types are harmonized in ASQ into intuitive evidence groups making triangulation of evidence in different groups more accessible, without either the need to navigate to various area-specific topics or the need to formulate complex queries. As we have demonstrated with our systematic analysis, the evidence retrieved by ASQ can be of high value and relevance to a wide range of researchers epidemiology and health science to assist the triangulation of evidence in their research. This is a generalisable approach that could be applied to a wider array of knowledge graphs and evidence sources to support the development of tools for rapid “semi-automated” (assisted) review of pre-prints.

Recent advances in deep learning modelling have contributed to a significant improvement in natural language processing, and ASQ applies our previous method development^19^ in combining sequence classification Transformer models with text vector embeddings for the harmonization of entities in different taxonomies. ASQ is able to combine the functionalities of parsing free text to generate structural claims with the harmonization of heterogeneous entities and evidence to enable claims to be mapped to with evidence both from literature and semantic knowledge as well as evidence from systematic association analysis. As part of the ASQ platform we developed a scoring mechanism to prioritise the retrieved evidence item, accounting for the semantic relevance of entities to the query of interest, as well as the strength of the evidence item per se. This score enables users to rapidly evaluate a wide range of evidence, whilst at the same time being able to assess the value of individual evidence items or evidence groups to the query to enable prioritisation.

On the other hand, it is worth pointing out that users should not be relying on metrics (whether they are ranking metrics, P-Values, or discrete categories of “accepting” / “inconclusive” / “rejecting”) as sole criteria when assessing evidence or as a substitute for detailed investigation, not just in ASQ but also interacting with data platforms. The nature of the heterogeneous source data means we strongly recommend users spend time investigating individual evidence sources using links provided by ASQ, as well understanding the various harmonization strategies in ASQ’s documentation, since data harmonization in itself is an opinionated way of data retrieval which might not be aligned with individual use cases. We seek to continue the development of ASQ in various aspects to improve the robustness of evidence retrieval and entity harmonization, as well as accessibility of evidence triangulation for researchers.

Whilst the ASQ platform offers a novel and accessible approach to querying a knowledge graph, there are some important limitations. The extraction of claims from a piece of text is rarely perfect, and genuine claims may be missed and others misinterpreted. The literature knowledge based represented by SemMedDB is also subject to the same limitations (using the same tool: SemRep), the triples extracted by SemRep are not context-specific (i.e. there is no information about which section of an abstract these come from, so may reflect hypothesis rather than conclusions) and in addition, literature evidence is subject to publication bias. The association evidence in EpiGraphDB is constrained to a subset of published GWAS datasets in OpenGWAS, and may omit important entities relating to claims in a query text. The ASQ approach should therefore be considered as a support tool that aids evidence identification to assess a claim, but not a comprehensive “fact-checker”.

## 4 Methods

### 4.1 Entity harmonization

Here we denote a **taxonomy** as a catalogue of terms in a specific domain, and an **ontology** as a tree representing the taxonomy terms in a hierarchical order (e.g. parent terms that are more generic versus descendant terms with more specific meanings; Figure 3B). In that sense, EFO is the ontology which ASQ uses to infer relationships between biomedical terms from the taxonomies of UMLS and GWAS traits. This is because EpiGraphDB incorporates semantic terms/triples in UMLS, but not its ontological hierarchies (since SemMedDB primarily curates derived triples with mechanistic predicates such as “CAUSES” but not comprehensive ontologies), and the GWAS traits are phenotypic trait names from genetic studies collated in the OpenGWAS platform. An **entity** is then defined as a member of a taxonomy, i.e. a biomedical concept can be represented in a taxonomy as one of its predefined members with an identifier and a label (e.g. UMLS term `C1305855` “Body mass index”) to various degrees of semantic affinity. Conceptually we refer to the process of resolving the mapping of terms from the claim triple with those from EpiGraphDB evidence as **entity harmonization**, as it harmonizes entities from different taxonomies into a unifying structure in the ontology (i.e. Figure 2). Our objective is to retrieve entities from EpiGraphDB that are semantically similar and ontologically meaningful with respect to the query terms, while ensuring broader relevant terms are retrieved by not restricting to identical token-level resemblance (which can also be achieved in ASQ by setting very high semantic similarity thresholds). To this end ASQ retrieves EFO entities that would sufficiently represent the query terms in the ontology hierarchy, then retrieve evidence entities that are semantically similar to the selected EFO entities.

In ASQ we measure the proximity between two entities in the semantic space by the **semantic similarity** of their labels, which is calculated as the cosine similarity ([0, 1]) between the text embedding vectors of the labels. Specifically, ontology terms from the `(Efo)` nodes, UMLS terms from the `(LiteratureTerm)` nodes, and GWAS traits from the `(Gwas)` nodes are pre-encoded by ScispaCy^20^ (`en_core_sci_lg-0.4.0`) into high-dimensional embedding vectors in an Elasticsearch vector store (Figure 1 left), which allows for fast retrieval of candidate entities via a k-nearest neighbour (kNN) search of the pre-computed vectors against the on-the-fly encoded vector of the query terms. On the one hand, entity representation via fast text embeddings is a naive approach on its own and candidate retrieval based on cosine similarity search can be highly sensitive to minor changes of the threshold, and on the other hand, sophisticated classification of entity relationships requires real-time inferencing between a large volume of candidate pairs using a dedicated classification model which is computationally resource-heavy. Thus for entity harmonization we implemented a two-stage approach that in the first stage the query terms are mapped to a handful of their close ontological representations in EFO, where simple semantic similarity measures are augmented by a dedicated ontology classification process (discussed below), and in the second stage greater number of evidence entities are retrieved for the corresponding EFO entities with kNN from the vector store. This enables robust and efficient retrieval of entities and evidence by ASQ.

The entity harmonization process starts with the retrieval of candidate EFO entities that semantically resemble the query terms, where ASQ attempts to select EFO entities that would qualify as either identical ontological representations of the query terms or as closely associated members in the hierarchy based on their **identity score** ([0, +∞)) with the query terms. The identity score is produced by BlueBERT-EFO^19^ which we trained on the term mappings of EFO-EFO terms and GWAS-EFO terms to infer the distance (number of steps/nodes) between a query term and an EFO term in the ontology tree. An identity score of 0 suggests the two terms are equivalent in the ontology, whereas a score of 1 suggests that the term of interest can be considered as either a direct parent term or a direct descendant term of the reference ontology term (in practice this can be relaxed to 1.5 as the inference model produces a *regression* score rather than a *classification* score) and scores above 2 suggest greater distance between the two terms. In previous research on the performance of entity retrieval by various methods^19^ we showed that BlueBERT-EFO as a task-specific bespoke model is able to retrieve candidate terms that are closer to a term of interest in the semantic rankings, than naive embeddings from general purpose models (e.g. ScispaCy, BioSentVec^21^, etc.). The retrieval of EFO candidates is also augmented with a pre-filtering step to remove ontology candidate terms that are overly generic to mitigate scenarios where retrieved evidence entities in subsequent steps are less relevant to query terms due to these evidence entities being mapped to generic ontology terms (such as an ontology term “disease”). This is done via the pre-computed **information content** (**IC**) **score** of EFO terms using a scaled Sanchez Information Content^22^ ([0, 1]) score where terms closer to an end node of the EFO tree have scores closer to 1 and terms closer to the origin have scores closer to 0. In both the interactive session and batch analysis mode, ASQ by default (identity score ≤ 1.5) seeks to select EFO candidates that would be either equivalent in the ontology to the query term, or a first-degree neighbour of it, as the basis for evidence identification, and in the interactive session users are able to further finetune the selection with the rest of the retrieved EFO candidates.

### 4.2 Evidence retrieval

As discussed in Section 2.1, epidemiological knowledge in EpiGraphDB is represented as two **evidence groups** in ASQ, i.e. the *triple and literature* group and the *association* group. From another perspective, the claim triple as well as evidence items in the two evidence groups are categorised by the **predicate group** according to whether for the claim or the evidence there is an indication of direction or not. Claims with predicates in the set “AFFECTS”, “CAUSES”, “PRODUCES” and “TREATS” parsed from SemRep are *directional* claims whereas predicates in the set “ASSOCIATED_WITH”, “COEXISTS_WITH”, and “INTERACTS_WITH” are *non-directional* claims. The set of included predicates are determined by the availability of corresponding predicates in the `(LiteratureTriple)` component of EpiGraphDB (Table 1). Similarly statistical results of association evidence are retrieved with predefined rules of predicate directions as well (Table 3). When the query claim is directional, directional results from `[MR_EVE_MR]` are retrieved as the basis of *supporting/reversal/insufficient* evidence types, and non-directional results from `[PRS]` and `[GEN_COR]` are retrieved as the *additional* evidence type. When the query claim is non-directional, results from all three sources are retrieved for the *supporting/insufficient* evidence types.

**Table 3.** Classification of retrieved evidence. Summary of how retrieved evidence items are classified based on the **predicate direction group, evidence group**, and **evidence type**. The notation *S* - *P* → *O* means a `Subject PREDICATE Object` triple where the predicate is directional (e.g. a “CAUSES” predicate versus a non-directional predicate “ASSOCIATED_WITH”) and the notation *S* - *P* → *O* means a triple with non-directional predicate.

|  | Supporting | Reversal | Insufficient | Additional |
| --- | --- | --- | --- | --- |
| Directional predicates |  |  |  |  |
| CAUSES, TREATS, PRODUCES, AFFECTS |  |  |  |  |
| Triple and literature | $S - P \rightarrow O$ | $O - P \rightarrow S$ | N/A | N/A |
| Association | $S - P \rightarrow O, P_p - Value < \pi$ | $O - P \rightarrow S, P_p - Value < \pi$ | $S - P \rightarrow O, P_p - Value \geq \pi$ | non-directional $S - P - O$ |
| Non-directional predicates |  |  |  |  |
| INTERACTS_WITH, COEXISTS_WITH, ASSOCIATED_WITH |  |  |  |  |
| Triple and literature | $S - P - O$ | N/A | N/A | N/A |
| Association | $S - P - O, P_p - Value < \pi$ | N/A | $S - P - O, P_p - Value \geq \pi$ | N/A |

The **evidence types** for each of the **evidence groups** and **predicate groups** are defined as below and summarised in Table 1:

- ***Supporting*** evidence items are those that provide sufficient evidence in support of the query claim. For *triple and literature* evidence, evidence from mapped literature terms which share the same predicate with the claim triple are *supporting* evidence. For *association* evidence, for *directional* claims, evidence from MR-EvE with strong statistical evidence (using P-Value as the parameter and default to P-Value ≤ 1*e* - 2) are identified as the *supporting* evidence, whereas for *non-directional* claims, evidence is identified from MR-EvE, PRS, and GEN_COR with strong statistical evidence.
- ***Reversal*** evidence items are those that could sufficiently contradict the claim with identified evidence from the reverse direction, and therefore is only applicable to *directional* predicates. In other words, for a claim “Obesity CAUSES Asthma” evidence that would support a claim “Asthma CAUSES Obesity” would be considered as a *reversal* evidence item, because it reverses the direction. For both *triple and literature* evidence group and *association* group, evidence item where its *source node* is a mapped *evidence object* entity and its *target node* is a mapped *evidence subject* entity is identified as a reversal evidence item. For *association* evidence the statistical threshold for supporting evidence also applies.
- ***Insufficient*** evidence items are identified as candidates for supporting evidence and reversal evidence (when applicable) which fail to meet the desired strength of evidence. This only applies to association evidence, for which P-Value is a quantitative measure. The aim of identifying insufficient evidence is to provide findings on the existence of systematic results, i.e. to determine whether the lack of evidence for a claim of interest is due to the absence of evidence (e.g. not curated by EpiGraphDB), or due to existing results failing to support/contradict a claim with sufficient strength.
- ***Additional*** evidence items are identified as evidence that could be of potential interest to users for further investigation, but which may not be sufficiently specific to inform the acceptance or rejection of a claim. For *association* evidence, when the claim is *directional*, non-directional evidence from PRS and GEN_COR are candidates for *additional* evidence.

### 4.3 Score metrics to measure retrieved evidence

We introduce scores for the retrieved evidence in order to facilitate the assessment of individual evidence items and provide a simple way to compare between evidence items and groups. However as naive assessment metrics they should be used for simple comparisons and should not replace the actual investigation into specific evidence details.

The mapping score *P*_mapping_ ([0, 1]; Equation 1) of retrieved evidence measures the overall deviation in terms of semantic similarity (*S*) between the retrieved evidence entities and the original query claim terms, which is a product of semantic similarity scores of associated entities in the entity harmonization stage. A high score indicates that the retrieved evidence is of high semantic proximity to the query claim of interest, whereas a low score suggests that the semantic relevance of the retrieved entity to the claim is low and therefore the relevance of the evidence to the query should be discounted by the low semantic relevance. If multiple *j* EFO entities are identified for a query term, but these map to the same evidence entity, the route with the highest score value is chosen as the basis for mapping score calculation. In addition, for triple entities the query terms are added as *pseudo*-ontology entities as they share the same UMLS taxonomy.

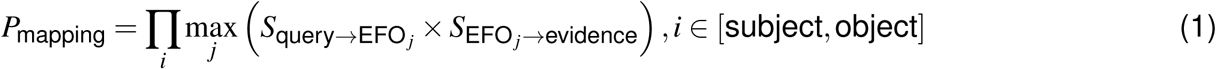

For *triple and literature* evidence, the strength of the evidence *P*_T&L._ ([1, +∞); Equation 2a) is measured by the number of source literature items containing the semantic triple evidence. The baseline for the evidence score is 1 where the semantic triple is associated with 1 source literature article. Therefore the *evidence score* for *triple and literature* evidence *E*_T&L._ ([0, +∞); Equation 2b) is a product of the evidence strength and mapping score, where in a typical scenario when there is an exact mapping (mapping score 1) of the involved entities and there is 1 source literature article the individual evidence item will have a baseline score of 1.

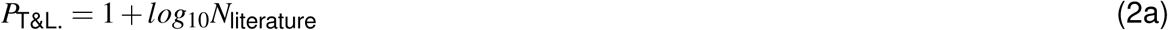

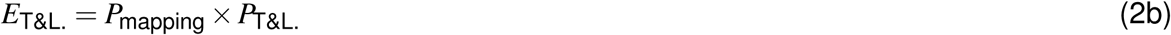

For *association* evidence, the strength of the evidence *P*_Assoc._ ([0, +∞); Equation 3a) is measured with the standardized effect size of the statistical results where a unit absolute standardized effect size produces a score of 1 as the baseline. Similarly the evidence score for *association* evidence *E*_Assoc._ ([0, +∞); Equation 3b) is the product of the mapping status and the association evidence strength.

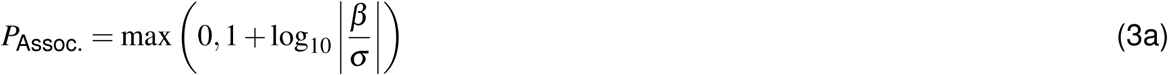

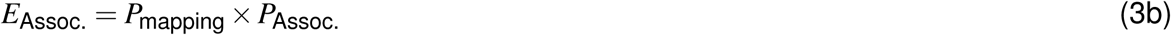

ASQ calculates the aggregate score and average score for each of the evidence groups and evidence types. Simple comparisons could be made (but should not be the substitute for further investigations) in cases such as between the supporting evidence group and reversal evidence group of a claim, as well as between two supporting evidence items. However comparisons between the supporting evidence group and the insufficient/additional evidence groups via quantitative measures are not appropriate as insufficient/additional evidence types by definition do not assess the query claim by metrics, and should instead be interpreted by the user based on their own knowledge. Similarly comparisons between the *triple and literature* group and the *association* group by metrics is not appropriate as they do not share a common measurement unit for their scores.

## Data Availability

All data produced are available online as part of the code repository https://github.com/mrcieu/epigraphdb-asq.

## Code availability

Source code for the ASQ platform and relevant analysis scripts can be found via https://github.com/mrcieu/epigraphdb-asq. Tutorial on programmatically accessing the ASQ platform can be found via this Jupyter notebook https://github.com/MRCIEU/epigraphdb-asq/blob/master/analysis/notebooks/programmatic-access.ipynb.

