## supplementary materials for "Triangulating evidence in health sciences with Annotated Semantic Queries"

### 1 Supplementary tables

**Supplementary Table 1. EpiGraphDB knowledge on UMLS terms and relationships**
Summary counts of UMLS terms (top) and UMLS relationships (bottom) by their respective categories covered in this study. The UMLS terms and predicates are integrated as `(LiteratureTerm)` and `(LiteratureTriple)` knowledge in EpiGraphDB (version 1.0). Query text is parsed into a set of UMLS triples consisting of UMLS terms as subjects and objects, and UMLS relationships as predicates. In addition, the triple evidence retrieved also consists of UMLS triples as available from EpiGraphDB. **Top**: UMLS term (e.g. `C0424678` "Lean body mass") counts grouped by their semantic types, where the description of each semantic type is obtained from[1]. **Bottom**: UMLS semantic network relationships[2] as covered in this study, where "predicate group" refers to the harmonized predicate group as discussed in Section 4.1.

| UMLS terms | | |
|---|---|---|
| **Semantic type** | **Description** | **Count** |
| `gngm` | Gene or Gnome | 42,675 |
| `aapp` | Amino Acid, Peptide, or Protein | 34,713 |
| `orch` | Organic Chemical | 31,136 |
| `phsu` | Pharmacologic Substance | 15,192 |
| `dsyn` | Disease or Syndrome | 14,912 |
| `clnd` | Clinical Drug | 1,638 |
| `inch` | Inorganic Chemical | 1,584 |
| `hops` | Hazardous or Poisonous Substance | 1,555 |
| `enzy` | Enzyme | 1,036 |
| `horm` | Hormone | 823 |
| `clna` | Clinical Attribute | 342 |
| `chem` | Chemical | 9 |
| UMLS predicates | | |
| **Predicate** | **Predicate group** | **Count** |
| `INTERACTS_WITH` | Undirectional | 1,272,318 |
| `COEXISTS_WITH` | Undirectional | 892,955 |
| `ASSOCIATED_WITH` | Undirectional | 302,338 |
| `CAUSES` | Directional | 200,813 |
| `TREATS` | Directional | 191,858 |
| `PRODUCES` | Directional | 179,281 |
| `AFFECTS` | Directional | 128,500 |

### Supplementary Table 2. Knowledge triple and literature evidence

Summary of triple and literature evidence counts by top triple groups. A triple evidence item takes the form of a UMLS triple e.g. "Obesity CAUSES Asthma" where the subject term *"Obesity"* (UMLS ID `C0028754`) and the object term *"Asthma"* (UMLS ID `C0004096`) both have type `dsyn` (Disease). For triple evidence items, we calculated the summary counts grouped by their predicate term, as well as the semantic types of subject and object entities, and then extracted the top 5 (by triple count) entries within each predicate group. We then calculated the number of literature evidence items associated with entry. Descriptions of the subject/object semantic types are available in Supplementary Table 1.

| Predicate group | Subject type | Object type | Triple count | Literature count |
|---|---|---|---|---|
| AFFECTS | aapp | dsyn | 37,243 | 57,928 |
| AFFECTS | gngm | dsyn | 37,243 | 57,928 |
| AFFECTS | dsyn | dsyn | 29,167 | 58,753 |
| AFFECTS | orch | dsyn | 17,038 | 25,202 |
| AFFECTS | phsu | dsyn | 17,038 | 25,202 |
| ASSOCIATED_WITH | aapp | dsyn | 247,248 | 597,129 |
| ASSOCIATED_WITH | gngm | dsyn | 247,248 | 597,129 |
| ASSOCIATED_WITH | phsu | dsyn | 29,425 | 111,438 |
| ASSOCIATED_WITH | enzy | dsyn | 28,862 | 61,964 |
| CAUSES | dsyn | dsyn | 85,231 | 222,462 |
| CAUSES | aapp | dsyn | 49,178 | 125,153 |
| CAUSES | gngm | dsyn | 49,178 | 125,153 |
| CAUSES | orch | dsyn | 19,064 | 46,666 |
| CAUSES | phsu | dsyn | 19,064 | 71,138 |
| COEXISTS_WITH | aapp | aapp | 343,575 | 521,867 |
| COEXISTS_WITH | gngm | gngm | 343,575 | 521,867 |
| COEXISTS_WITH | aapp | gngm | 343,575 | 521,867 |
| COEXISTS_WITH | gngm | aapp | 343,575 | 521,867 |
| COEXISTS_WITH | dsyn | dsyn | 150,166 | 385,349 |
| INTERACTS_WITH | gngm | gngm | 694,873 | 1268,896 |
| INTERACTS_WITH | gngm | aapp | 682,155 | 1268,896 |
| INTERACTS_WITH | aapp | gngm | 675,239 | 1246,995 |
| INTERACTS_WITH | aapp | aapp | 662,521 | 1246,995 |
| INTERACTS_WITH | aapp | phsu | 126,849 | 221,742 |
| PRODUCES | aapp | aapp | 111,929 | 155,703 |
| PRODUCES | aapp | gngm | 111,929 | 155,703 |
| PRODUCES | gngm | gngm | 111,929 | 155,703 |
| PRODUCES | gngm | aapp | 111,929 | 155,703 |
| PRODUCES | aapp | phsu | 12,706 | 26,122 |
| TREATS | phsu | dsyn | 150,033 | 632,300 |
| TREATS | orch | dsyn | 82,263 | 274,589 |
| TREATS | gngm | dsyn | 20,354 | 119,075 |
| TREATS | aapp | dsyn | 20,354 | 119,075 |

### Supplementary Table 3. Association evidence

Summary of association evidence counts by association type and the category of the involved GWAS. An association evidence item is a quantitative relationship between two GWAS traits which comes from three EpiGraphDB relationship sources `[MR_EVE_MR]`, `[GEN_COR]`, `[PRS]`. For `GEN_COR` and `PRS` items we grouped them by the sub categories (e.g. `ukb-b`), and for `MR_EVE_MR` items due to the high pairwise density nature of the `[MR_EVE_MR]` relationship we grouped them by the broad categories (e.g. `ukb`) and extracted the top 5 entries to be succinct. Cells in column *GWAS categories* show the categories which the source and target GWAS-es belong to (e.g. for [`ukb-a`, `ukb-b`] it could be the source GWAS belongs to `ukb-a` and the target GWAS belongs to `ukb-b`, or the other way around). Descriptions on the GWAS categories are available from OpenGWAS documentation[3].

| Association type | GWAS categories | Evidence count |
|---|---|---|
| GEN_COR | [ukb-b, ukb-b] | 453,752 |
| GEN_COR | [ukb-a, ukb-b] | 286,536 |
| GEN_COR | [ukb-a, ukb-a] | 180,536 |
| GEN_COR | [ukb-b, ukb-d] | 133,554 |
| GEN_COR | [ukb-a, ukb-d] | 84,266 |
| GEN_COR | [ukb-d, ukb-d] | 38,908 |
| PRS | [ieu-a, ukb-a] | 70,926 |
| PRS | [ukb-b, ieu-a] | 45,394 |
| PRS | [ukb-a, ukb-a] | 2,198 |
| PRS | [ukb-b, ukb-a] | 704 |
| MR_EVE_MR | [ukb, ukb] | 8,966,440 |
| MR_EVE_MR | [prot, ukb] | 5,028,904 |
| MR_EVE_MR | [ubm, ukb] | 3,833,948 |
| MR_EVE_MR | [prot, prot] | 3,109,406 |
| MR_EVE_MR | [prot, ubm] | 1,974,611 |

**Supplementary Table 4. Notation conventions**

Notation for source biomedical entities and knowledge in EpiGraphDB. The graph database of EpiGraphDB models biomedical entities as `(nodes)` and their relationships as `[RELATIONSHIPS]`, where we refer to these sources in the context of the study using Neo4j Cypher syntax.

| Graph elements | Label | Description |
|---|---|---|
| | | EpiGraphDB nodes |
| `(Efo)` | EFO | Experimental Factor Ontology[4] which is widely used in categorising GWAS traits. |
| `(LiteratureTerm)` | literature terms | UMLS Metathesaurus terms describing `(LiteratureTriple)`s curated in SemMedDB[5]. The subject and object entities of *triple and literature* evidence in ASQ. |
| `(LiteratureTriple)` | literature triple | Literature-derived semantic triples curated in SemMedDB. The semantic triple components of *triple and literature* evidence in ASQ. |
| `(Literature)` | source literature | PubMed[6] literature from which `(LiteratureTriple)`s are derived. The literature components of *triple and literature* evidence in ASQ. |
| `(Gwas)` | GWAS / GWAS traits | Traits of genome-wide association studies curated in the OpenGWAS consortium[3]. OpenGWAS curates the various GWAS set used in the association relationships below. The subject and object entities of *association* evidence in ASQ. |
| | | EpiGraphDB relationships |
| `[MR_EVE_MR]` | MR-EvE[7] | Systematic analysis results of pairwise causal effects using Mendelian Randomization between two GWAS in MR-Base[8] and OpenGWAS[3]. Constituent source evidence for *association* evidence in ASQ. |
| `[PRS]` | PRS / PRS Atlas[9] | Systematic analysis results of pairwise polygenic risk score associations between two GWAS in MR-Base and UKBiobank GWAS. Constituent source evidence for *association* evidence in ASQ. |
| `[GEN_COR]` | GEN_COR[10] | Systematic analysis results of pairwise genetic correlations of the UKBiobank GWAS. Constituent source evidence for *association* evidence in ASQ. |

**Supplementary Table 5. Systematic analysis: parametric configuration**

Configurable parameters in ASQ in the processes of entity harmonization and evidence retrieval, and the specific values used in the systematic analysis results in Section 2.2. For conceptual discussions and technical details on the parameters please refer to Section 4. Further documentation on the ASQ platform can be found at https://asq.epigraphdb.org/docs.

| Parameter | Description | Value |
|---|---|---|
| Semantic similarity threshold for ontology candidates | Semantic similarity between terms is calculated as the cosine similarity between two vectors of encoded terms via a text embedding model (ScispaCy[11]). The threshold is the primary metric in determining which ontology entities are retrieved as candidates in the entity harmonization. | 0.7 |
| Number of retrieved ontology candidates | Maximum number of candidates to retrieve from the vector store that are above the semantic similarity threshold. | 10 |
| Information content score threshold | The information content score measures the concreteness of a node in the ontology tree, with higher values associated with nodes towards the end of the branch. For example, for cancer related terms, the term "carcinoma" is roughly at 0.6. Candidates below the threshold will be removed, which is to mitigate scenarios where evidence entities retrieved in subsequent stages are of low relevancy to the query term due to them being mapped to a generic ontology term (e.g. "disease"). | 0.6 |
| Identity score threshold | The identity score measures the relationship between the query term and the reference ontology in the ontology space, i.e. they are identical (closer to 0), a direct parent-descendent pair (closer to 1), relationship of further distance (greater than 2). The threshold determines which retrieved ontology *candidates* qualify as the associated ontology *entities* of the query term. | 1.5 |
| Semantic similarity threshold for evidence entities | Threshold for retrieving evidence (UMLS and GWAS trait) entities. | 0.7 |
| Number of retrieved evidence entities | Maximum number of retrieved evidence entities. | 20 |
| Statistical significance threshold | P-Value threshold to categorise association evidence items to the evidence types of "supporting", "reversal", and "insufficient". By default ASQ will seek to identify evidence items that quantitatively qualify at a sufficient statistically significant level but this behaviour can be overridden by the user to other levels. | 1e-2 |

**Supplementary Table 6. Systematic analysis: summary statistics**

Entity count on claim triples, retrieved entities and retrieved evidence in the systematic analysis results. For the *claim triples* column the main value reports the number of terms (subjects and objects) in claim triples that are identified to contain associated entities and evidence in ASQ, and the value in parentheses reports the number of terms in the initial claim parsing sample. For the *EFO entities*, *UMLS entities*, and *trait entities* columns the main value reports the number of retrieved entities for the claim triples, and the value in parentheses reports the number of entities with semantic similarity scores above 0.85 (as a conventional threshold to signify entities that are similar to the claim terms). For the *T&L. evidence* (triple and literature evidence) and *Assoc. evidence* (association evidence) columns the main value reports the number of evidence items in the group across all evidence types, and the values in parentheses report the number of supporting evidence items and the number of supporting evidence items with evidence scores above 1.

| Predicate | Claim triples | EFO entities | UMLS entities | Trait entities | T&L. evidence | Assoc. evidence |
|---|---|---|---|---|---|---|
| Directional predicates | | | | | | |
| `AFFECTS` | 85 | 344 | 7,984 | 1,432 | 1,955 | 7,735 |
| | (2,487) | (288) | (3,806) | (679) | (375, 141) | (372, 196) |
| `CAUSES` | 67 | 298 | 7,824 | 1,280 | 1,541 | 6,097 |
| | (1,127) | (242) | (3,797) | (385) | (1,266, 482) | (412, 123) |
| `TREATS` | 21 | 70 | 2,200 | 116 | 483 | 1,911 |
| | (5,779) | (61) | (1,077) | (100) | (283, 143) | (83, 55) |
| Non-directional predicates | | | | | | |
| `ASSOCIATED_WITH` | 66 | 224 | 5,316 | 2,254 | 660 | 2,442 |
| | (1,722) | (207) | (2,560) | (474) | (211, 73) | (3,106, 1,064) |
| `COEXISTS_WITH` | 170 | 744 | 18,430 | 4,000 | 1,700 | 6,290 |
| | (2,712) | (681) | (8,131) | (1,402) | (1,820, 736) | (2,401, 1,185) |
| `INTERACTS_WITH` | 4 | 10 | 354 | 34 | 40 | 148 |
| | (609) | (8) | (136) | (30) | (4, 1) | (46, 17) |

**Supplementary Table 7. Entity harmonization stage: distribution of query UMLS entities and EFO entities by UMLS semantic type**

Distribution of the identified query UMLS entities and harmonized EFO entities in the systematic analysis, ordered descendingly by EFO entity count to be in line with Figure 5 and Supplementary Figure 1. The query UMLS entities are derived from parsing the MedRxiv abstracts from 2020-01-01 to 2021-12-31, and the EFO entities are retrieved from EpiGraphDB in the process of entity harmonization of the corresponding query UMLS entities.

| Semantic type | Description | Query UMLS entity count | EFO entity count |
|---|---|---|---|
| `dsyn` | Disease or Syndrome | 888 | 7,831 |
| `mobd` | Mental or Behavioral Dysfunction | 125 | 1,708 |
| `neop` | Neoplastic Process | 138 | 1,516 |
| `sosy` | Sign or Symptom | 124 | 562 |
| `phsu` | Pharmacologic Substance | 447 | 488 |
| `bacs` | Biologically Active Substance | 98 | 314 |
| `patf` | Pathologic Function | 171 | 303 |
| `fndg` | Finding | 509 | 266 |
| `orch` | Organic Chemical | 253 | 223 |
| `orgf` | Organism Function | 74 | 190 |
| `aapp` | Amino Acid, Peptide, or Protein | 487 | 152 |
| `bpoc` | Body Part, Organ, or Organ Component | 20 | 129 |
| `horm` | Hormone | 31 | 106 |
| `orga` | Organism Attribute | 31 | 77 |
| `bhvr` | Behavior | 2 | 76 |
| `topp` | Therapeutic or Preventive Procedure | 509 | 60 |
| `ortf` | Organ or Tissue Function | 74 | 47 |
| `phsf` | Physiologic Function | 44 | 42 |
| `anab` | Anatomical Abnormality | 9 | 30 |
| `clna` | Clinical Attribute | 24 | 30 |
| `gngm` | Gene or Genome | 504 | 20 |
| `medd` | Medical Device | 79 | 10 |
| `hops` | Hazardous or Poisonous Substance | 26 | 8 |
| `cell` | Cell | 49 | 8 |
| `inpo` | Injury or Positioning | 45 | 5 |

**Supplementary Table 8. Evidence retrieval stage: summary statistics**

Summary of evidence scores and constituent scores for retrieved evidence in the systematic analysis, categorised by predicate group, predicate term, and evidence type. A score metric is reported as "aggregated value (average value)", i.e. there are 372 retrieved association evidence items in the supporting evidence type with a predicate "AFFECTS", with an aggregated score of 387.40 and an average score of 1.04 per item.

| Predicate group | Predicate term | Evidence type | Item count | Strength score | Mapping score | Evidence score |
|---|---|---|---|---|---|---|
| | | Association evidence | | | | |
| **Directional** | **AFFECTS** | **Supporting** | 372 | 676.04 (1.82) | 212.94 (0.57) | 387.40 (1.04) |
| | | Reversal | 343 | 634.07 (1.85) | 206.75 (0.60) | 382.35 (1.11) |
| | | Insufficient | 3,139 | 2,566.68 (0.82) | 1,870.44 (0.60) | 1,532.54 (0.49) |
| | | Additional | 289 | 322.96 (1.12) | 166.20 (0.58) | 183.41 (0.63) |
| | **CAUSES** | **Supporting** | 412 | 719.92 (1.75) | 204.93 (0.50) | 361.96 (0.88) |
| | | Reversal | 399 | 700.80 (1.76) | 199.85 (0.50) | 354.40 (0.89) |
| | | Insufficient | 2,732 | 2,416.45 (0.88) | 1,354.39 (0.50) | 1,189.06 (0.44) |
| | | Additional | 1,111 | 2,127.96 (1.92) | 499.12 (0.45) | 928.82 (0.84) |
| | **TREATS** | **Supporting** | 83 | 158.41 (1.91) | 47.03 (0.57) | 89.99 (1.08) |
| | | Reversal | 166 | 328.81 (1.98) | 97.86 (0.59) | 194.38 (1.17) |
| | | Insufficient | 111 | 100.73 (0.91) | 57.31 (0.52) | 52.16 (0.47) |
| | | Additional | 0 | 0.00 (N/A) | 0.00 (N/A) | 0.00 (N/A) |
| **Non-directional** | **ASSOCIATED_WITH** | **Supporting** | 3,106 | 5,814.56 (1.87) | 1,544.26 (0.50) | 2,872.33 (0.92) |
| | | Insufficient | 5,272 | 4,570.48 (0.87) | 2,839.05 (0.54) | 2,441.49 (0.46) |
| | **COEXISTS_WITH** | **Supporting** | 2,401 | 4,409.50 (1.84) | 1,373.44 (0.57) | 2,519.11 (1.05) |
| | | Insufficient | 7,315 | 6,202.52 (0.85) | 4,261.65 (0.58) | 3,623.38 (0.50) |
| | **INTERACTS_WITH** | **Supporting** | 46 | 76.98 (1.67) | 27.33 (0.59) | 45.98 (1.00) |
| | | Insufficient | 174 | 157.41 (0.90) | 102.48 (0.59) | 92.98 (0.53) |
| | | Triples & literature evidence | | | | |
| **Directional** | **AFFECTS** | **Supporting** | 375 | 497.90 (1.33) | 273.09 (0.73) | 367.50 (0.98) |
| | | Reversal | 291 | 385.62 (1.33) | 211.45 (0.73) | 285.21 (0.98) |
| | **CAUSES** | **Supporting** | 1,266 | 1,779.61 (1.41) | 875.66 (0.69) | 1,244.00 (0.98) |
| | | Reversal | 954 | 1,279.71 (1.34) | 632.01 (0.66) | 847.10 (0.89) |
| | **TREATS** | **Supporting** | 283 | 439.03 (1.55) | 191.45 (0.68) | 303.71 (1.07) |
| | | Reversal | 0 | 0.00 (N/A) | 0.00 (N/A) | 0.00 (N/A) |
| **Non-directional** | **ASSOCIATED_WITH** | **Supporting** | 211 | 286.43 (1.36) | 149.63 (0.71) | 205.82 (0.98) |
| | **COEXISTS_WITH** | **Supporting** | 1,820 | 2,638.37 (1.45) | 1,269.99 (0.70) | 1,857.41 (1.02) |
| | **INTERACTS_WITH** | Supporting | 4 | 5.81 (1.45) | 2.68 (0.67) | 4.08 (1.02) |

**Supplementary Table 9. Systematic analysis results: top claim triples by retrieved evidence**

Top claim triples sorted by number of source claim abstract documents and the combined score between supporting triple and literature evidence as well as association evidence. For example, for claim "Coronavirus infections CAUSES Disease" there are 9 abstracts from MedRxiv articles associated with this claim, and EpiGraphDB-ASQ identifies 17 supporting triple and literature evidence ("T&L: S") items with an aggregated score of 21.19, and no supporting association evidence items ("Assoc: S"). In addition, for the reversal evidence to the claim (i.e. evidence supporting a claim "Disease CAUSES Coronavirus infections"), EpiGraphDB-ASQ identifies 8 reversal triple and literature evidence items ("T&L: R") with an aggregated score of 8.25, and 2 reversal association evidence items ("Assoc: R") with an aggregated score of 1.65. Reversal evidence is not applicable for "non-directional" claims without indication of direction in the predicate relationship. Interactive results on retrieved evidence for all claims in the systematic analysis are available on `https://asq.epigraphdb.org/medrxiv-analysis`.

| Claim triple | Lit. | T&L: S | T&L: R | Assoc: S | Assoc: R |
|---|---|---|---|---|---|
| **Directional predicates** | | | | | |
| Coronavirus Infections CAUSES Disease | 9 | 21.19 (17) | 8.25 (8) | 0.00 (0) | 1.65 (2) |
| Blood Glucose AFFECTS Diabetes Mellitus, Non-Insulin-Dependent | 2 | 3.89 (4) | 2.49 (3) | 9.55 (10) | 9.55 (9) |
| Diabetes Mellitus, Non-Insulin-Dependent AFFECTS Parkinson Disease | 2 | 4.61 (4) | 5.77 (6) | 1.97 (2) | 0.96 (1) |
| Low Back Pain CAUSES Chronic pain | 1 | 2.42 (4) | 2.42 (4) | 156.09 (222) | 162.14 (225) |
| Valvular disease CAUSES Heart failure | 1 | 74.78 (74) | 41.35 (49) | 32.68 (32) | 25.39 (24) |
| Metabolic Diseases CAUSES Liver diseases | 1 | 50.48 (57) | 50.10 (58) | 2.22 (2) | 0.00 (0) |
| Heart Diseases CAUSES Pulmonary Hypertension | 1 | 39.48 (35) | 34.72 (39) | 6.20 (8) | 2.84 (3) |
| Myocardial Infarction CAUSES Acute myocardial infarction | 1 | 11.67 (11) | 10.76 (10) | 31.11 (20) | 29.84 (20) |
| **Non-directional predicates** | | | | | |
| Triglycerides COEXISTS_WITH Very low density lipoprotein | 2 | 42.05 (47) | N/A | 60.40 (54) | N/A |
| Depressive disorder COEXISTS_WITH Parkinson Disease | 2 | 25.17 (24) | N/A | 6.09 (5) | N/A |
| Hepatic impairment COEXISTS_WITH Disease | 2 | 27.93 (27) | N/A | 0.93 (1) | N/A |
| Chronic disease COEXISTS_WITH Obesity | 2 | 17.80 (13) | N/A | 1.04 (1) | N/A |
| Disease COEXISTS_WITH Diabetes | 2 | 14.04 (11) | N/A | 2.23 (3) | N/A |
| Fatigue ASSOCIATED_WITH Disease | 2 | 0.80 (1) | N/A | 15.14 (21) | N/A |
| Sleep ASSOCIATED_WITH Alzheimer's Disease | 2 | 2.29 (3) | N/A | 11.18 (8) | N/A |
| Malignant Neoplasms COEXISTS_WITH Disease | 2 | 4.90 (5) | N/A | 6.17 (6) | N/A |

### 2 Supplementary figures

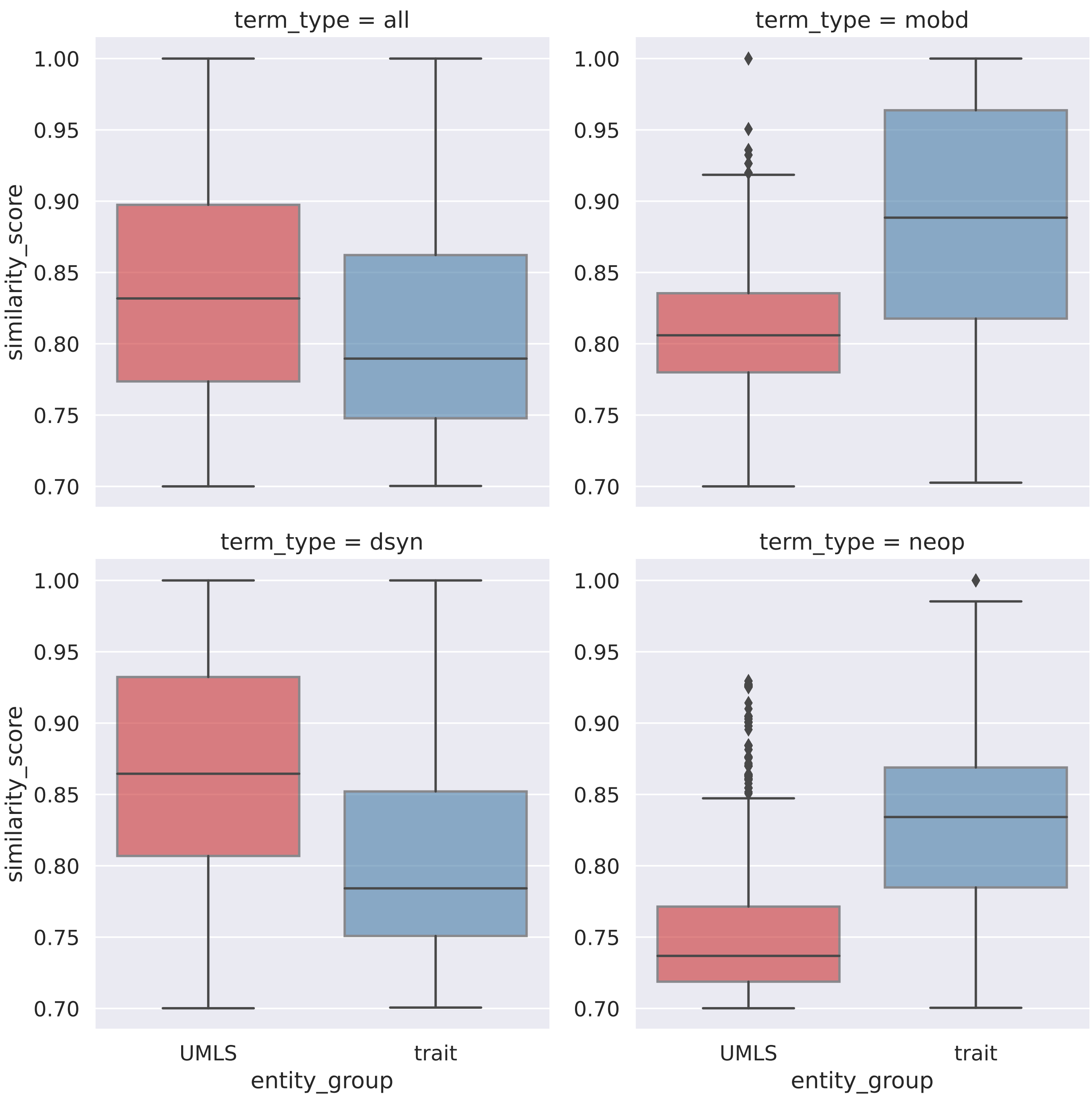


**Supplementary Figure 1. Entity harmonization: distribution of score metrics of retrieved UMLS and trait entities**
Distribution of semantic similarity scores as metrics used for retrieved UMLS and trait entities in the process of mapping with EFO entities, categorised by the semantic type of the initial query UMLS terms where the EFO entities are mapped to. This figure reports distributions in the top 3 semantic type groups by entity count (Supplementary Table 7 reports entity counts of all semantic types).

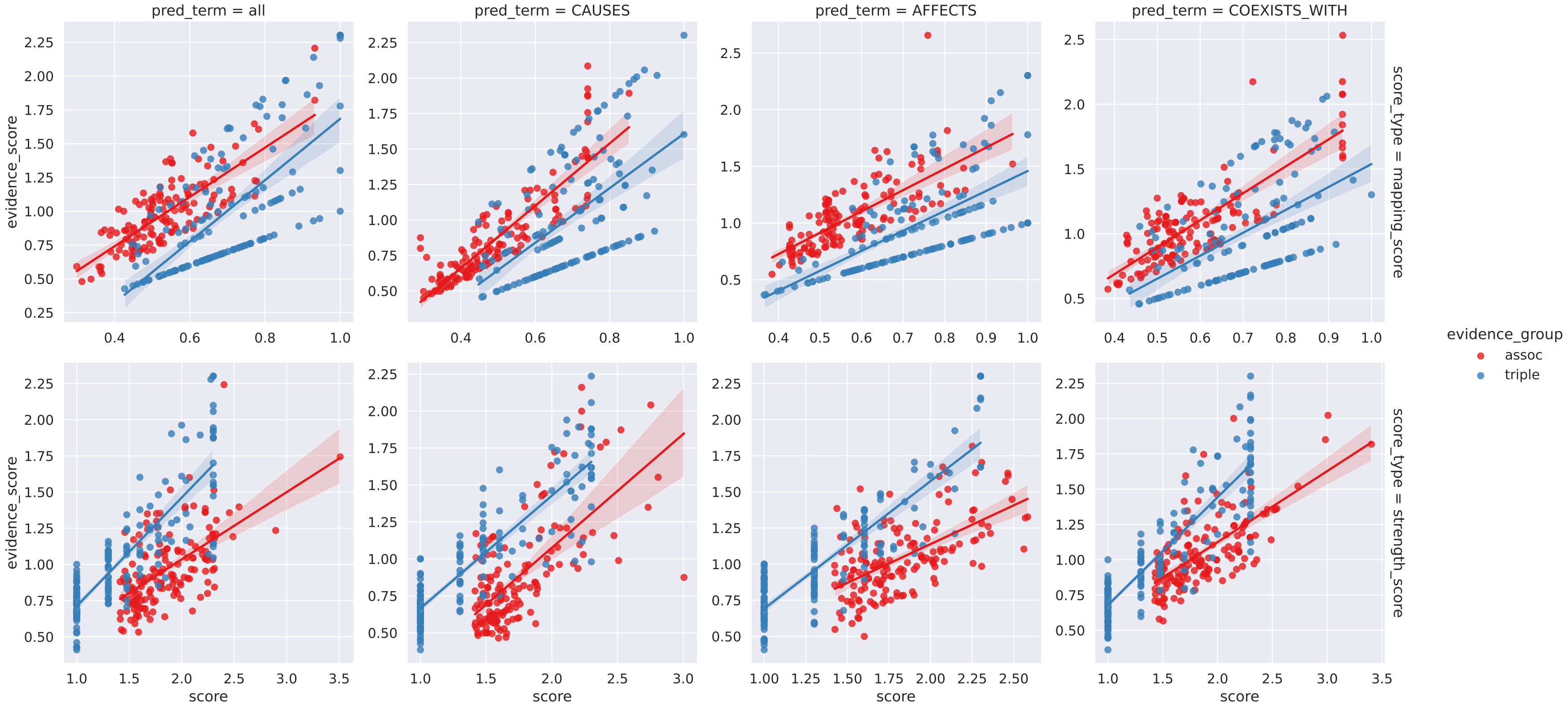


**Supplementary Figure 2. Evidence retrieval stage: relationship between evidence scores with its constituent scores (supporting evidence)**
The scatter plots report the relationship between evidence scores and mappings scores (**top**), as well as between evidence scores and strength scores (**bottom**). Point colours correspond to evidence groups of triple and literature group ("triple") and association group ("assoc").

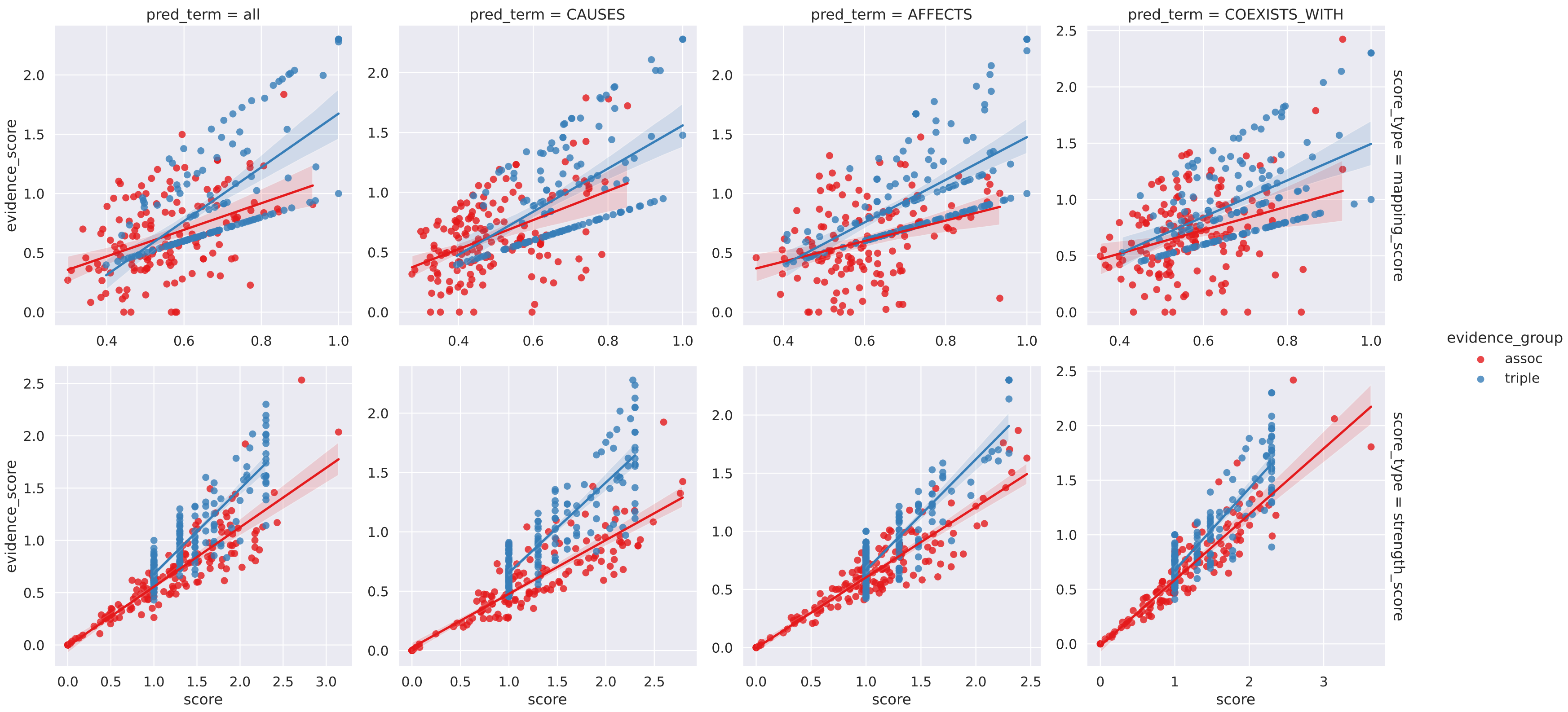


**Supplementary Figure 3. Evidence retrieval: relationship between evidence score with its constituent scores (all evidence types)**
The scatter plots report the relationship between evidence scores and mappings scores (**top**), as well as between evidence scores and strength scores (**bottom**). Point colours correspond to evidence groups of triple and literature group ("triple") and association group ("assoc").

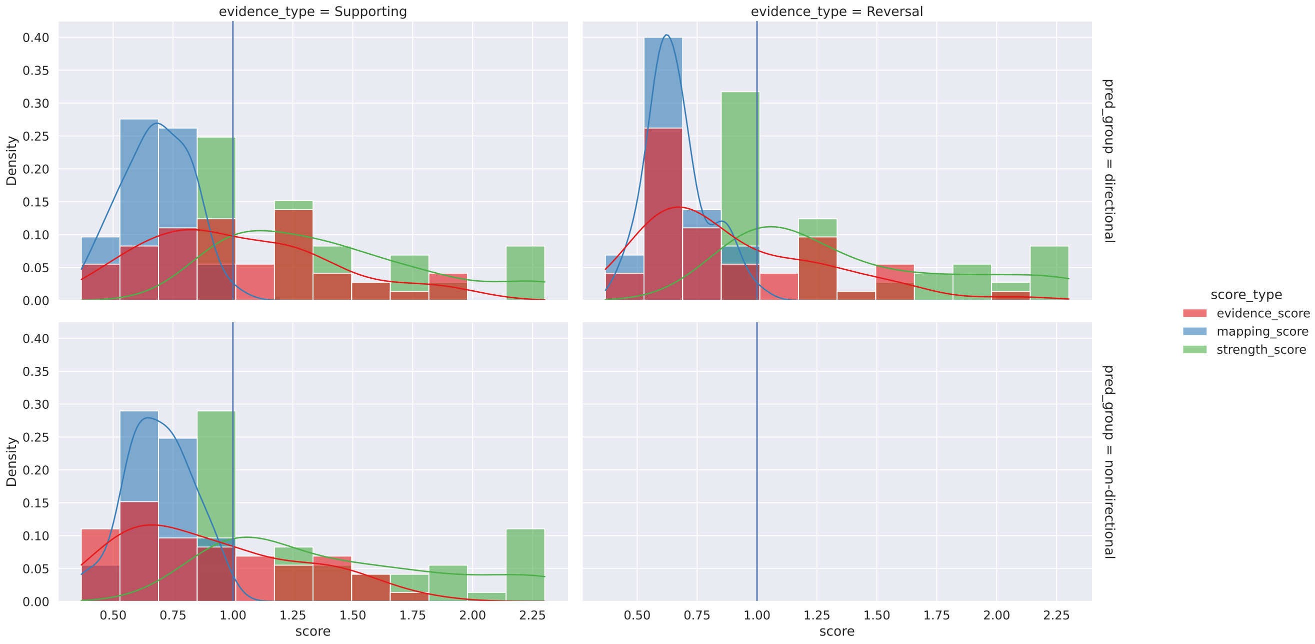


**Supplementary Figure 4. Evidence retrieval stage: distribution of evidence scores and constituent scores (triple and literature evidence group)**

Distribution of evidence scores and its constituent scores (entity mapping scores and evidence strength scores), for all evidence types (by columns) in the triple and literature evidence group and by predicate groups (by rows).

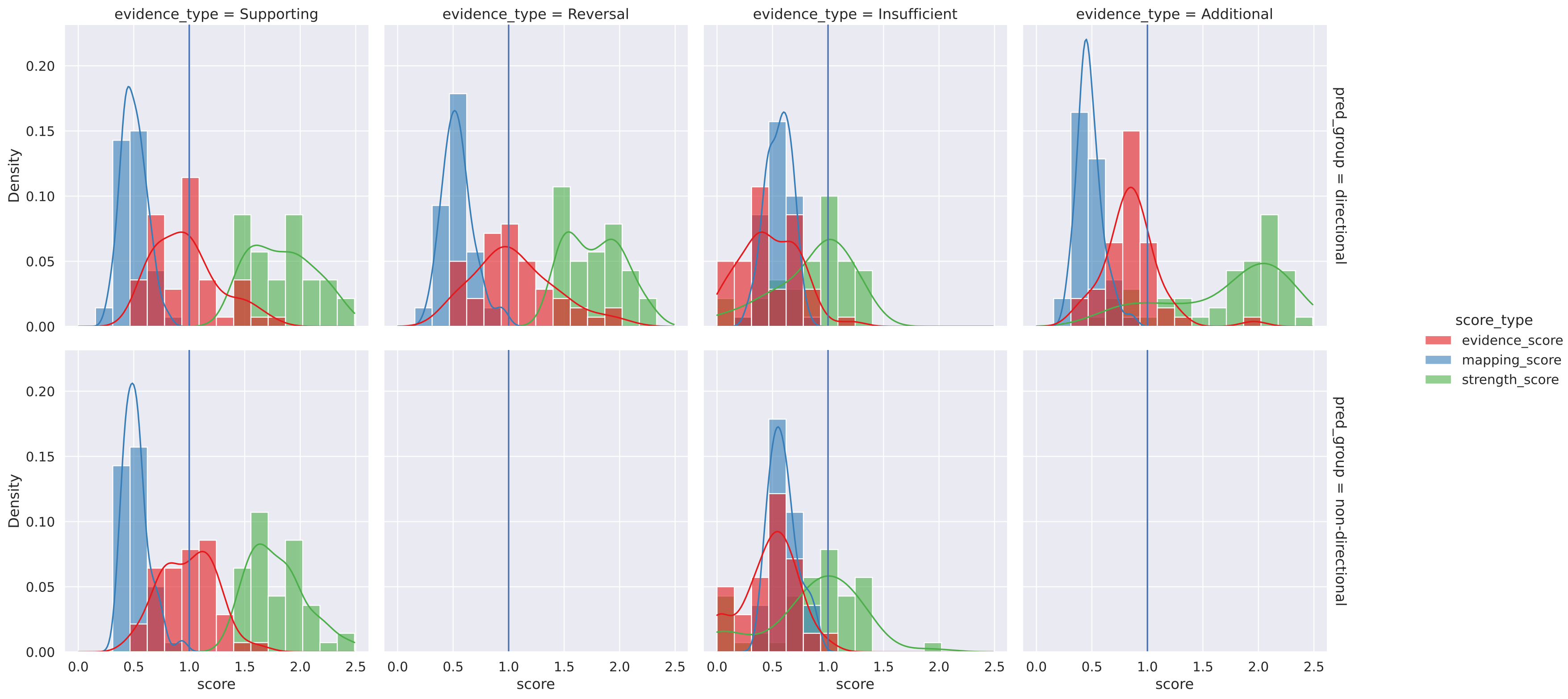


**Supplementary Figure 5. Evidence retrieval stage: distribution of evidence scores and constituent scores (associations evidence group)**
Distribution of evidence scores and its constituent scores (entity mapping scores and evidence strength scores), for all evidence types (by columns) in the association evidence group and by predicate groups (by rows).
